# Cell type-specific immune dysregulation in severely ill COVID-19 patients

**DOI:** 10.1101/2020.07.23.20161182

**Authors:** Changfu Yao, Stephanie A Bora, Tanyalak Parimon, Tanzira Zaman, Oren A Friedman, Joseph A Palatinus, Nirmala S Surapaneni, Yuri P Matusov, Giuliana Cerro Chiang, Alexander G Kassar, Nayan Patel, Chelsi ER Green, Adam W Aziz, Harshpreet Suri, Jo Suda, Andres A Lopez, Gislaine A Martins, Barry R Stripp, Sina A Gharib, Helen S Goodridge, Peter Chen

## Abstract

Coronavirus disease 2019 (COVID-19) has quickly become the most serious pandemic since the 1918 flu pandemic. In extreme situations, patients develop a dysregulated inflammatory lung injury called acute respiratory distress syndrome (ARDS) that causes progressive respiratory failure requiring mechanical ventilatory support. Recent studies have demonstrated immunologic dysfunction in severely ill COVID-19 patients. To further delineate the dysregulated immune response driving more severe clinical course from SARS-CoV-2 infection, we used single-cell RNA sequencing (scRNAseq) to analyze the transcriptome of peripheral blood mononuclear cells (PBMC) from hospitalized COVID-19 patients having mild disease (n = 5), developing ARDS (n = 6), and recovering from ARDS (n = 6). Our data demonstrated an overwhelming inflammatory response with select immunodeficiencies within various immune populations in ARDS patients. Specifically, their monocytes had defects in antigen presentation and deficiencies in interferon responsiveness that contrasted the higher interferon signals in lymphocytes. Furthermore, cytotoxic activity was suppressed in both NK and CD8 lymphocytes whereas B cell activation was deficient, which is consistent with the delayed viral clearance in severely ill COVID-19 patients. Finally, we identified altered signaling pathways in the severe group that suggests immunosenescence and immunometabolic changes could be contributing to the dysfunctional immune response. Our study demonstrates that COVID-19 patients with ARDS have an immunologically distinct response when compared to those with a more innocuous disease course and show a state of immune imbalance in which deficiencies in both the innate and adaptive immune response may be contributing to a more severe disease course in COVID-19.

## Introduction

SARS-CoV-2 infection has quickly spread worldwide to cause the COVID-19 pandemic ^1^. Coronaviruses are single, positive-stranded RNA viruses that can infect a range of hosts. Some are known to cause seasonal, upper respiratory infections (i.e. common colds), but coronaviruses that cause severe lower respiratory infection have emerged, including those that cause severe acute respiratory syndrome (SARS), Middle Eastern respiratory syndrome (MERS), and now COVID-19 ^2, 3, 4^. SARS-CoV-2 has reached pandemic proportions and is likely to remain a world health emergency for the foreseeable future due to lack of a vaccine, limited treatments, and a high likelihood of recurrent outbreaks. The World Health Organization lists the primary symptoms of COVID-19 as fever, dry cough, and fatigue but also include other symptoms such as diarrhea, loss of taste and smell, and rashes. Those over 60 years of age and people with obesity, cardiovascular disease, and diabetes have the highest risk for severe COVID-19 ^5, 6^. Most COVID-19 patients have mild respiratory illness, however, about 20% become seriously ill and require hospitalization due to pneumonia ^7^. This can progress into acute respiratory distress syndrome (ARDS) and systemic inflammation referred to as ” cytokine storm” ^8^.

Instead of beneficial antiviral immunity in response to infection, severe COVID-19 is characterized by dysregulated immune responses that allows the virus to persist, causing lung damage, ARDS, and systemic inflammation ^9^. While mechanisms underlying SARS-CoV-2 evasion of antiviral immunity and pathogenic inflammation are not clear at this time, commonalities in the pathogenic response with this novel coronavirus and SARS-CoV-1 and MERS-CoV have become apparent ^8, 10^. Cells sense RNA viruses using endosomal and cytosolic pattern recognition receptors (PRRs) which signal through other mediators including TNF receptor-associated factors (TRAF) 3 and 6 to activate interferon regulatory factors (IRF) and NFκB, resulting in transcription of early antiviral type I interferons by resident alveolar macrophages (AMs) and epithelial cells in the lungs, which sets up an immune response that clears the virus and resolves inflammation ^11^. SARS-CoV-1, and likely SARS-CoV-2, inhibit multiple viral sensing PRRs and downstream signals, effectively blocking recognition of virus and early antiviral type I interferon, and initiating a dysregulated inflammatory cascade that can lead to ARDS and systemic inflammation ^12, 13, 14^. Moreover, transcriptomic analysis of PBMC from COVID-19 patients found upregulated pro-inflammatory pathways in monocytes and CD4 T cells, suggesting that the basic hallmarks of the ” cytokine storm” in COVID-19 parallel SARS and MERS ^15^. However, we are now also appreciating immunologic dysfunctions that may be causing a more severe disease course ^16, 17, 18^.

COVID-19 patients have higher circulating levels of IL-6, TNF-α, and CXCL10, particularly those with severe disease, and these ” early” cytokines were sustained weeks into infection suggesting an inability to resolve inflammation ^19, 20^. Adaptive immune cells recruited from nearby lymph nodes (via circulatory and lymphatic systems) can also contribute to pathogenic inflammation in the lung, particularly if polarized to Th1 and Th17 responses that contribute to neutrophil recruitment and pro-inflammatory monocyte/macrophage activation ^21^. However, severe lung damage due to pneumonia or sepsis is more often characterized by a lack of adaptive immune cells in the periphery ^22, 23^. This is due to migration of cells to sites of inflammation, but also T cell dysfunction and death ^23, 24, 25^. Prolonged antigen stimulation and pro-inflammatory cytokine exposure cause T cell exhaustion and apoptosis, leading to insufficient B cell activation and loss of the immune resolving functions of T and B cells ^26, 27, 28^. Despite exuberant innate immunity, lymphopenia has been observed in COVID-19 and correlates with poor disease outcome ^27, 29^. Characterization of T cells from COVID-19 patients found increased surface expression of exhaustion markers Tim-3 and PD-1, decreased expression of pathways involved in T cell expansion, and increased expression of apoptotic pathways ^15, 20, 29^. Therefore, the dysfunction in circulating T and B cells during ARDS may result in an inability to resolve inflammation and perpetuate the systemic inflammation caused by the cytokine storm.

The innate-driven pathogenic inflammation and suppressed adaptive immunity during ARDS in COVID-19 patients indicate dysfunction in immune regulation and the switch from innate to adaptive immunity. To further understand this dysregulated immune response that drives patients to have more severe illness from SARS-CoV-2 infection, we used single-cell RNA sequencing (scRNAseq) to analyze the transcriptome of PBMC collected within the first 5 days of hospitalization of COVID-19 patients with a milder course of disease (mild group) and those that developed ARDS (severe group). Moreover, we investigated the signals that drives lung inflammation in ARDS by comparing the severe group to those recovering from ARDS (recovering group). Having sufficient numbers from each patient group allowed for a deeper analysis of each immune population and evaluation of the functional changes that may determine the severity of illness, as well as delineation of novel pathways that are activated during the resolution of lung inflammation. Severely ill patients had an overall higher activation of inflammatory pathways in comparison to mild and recovering patients. However, we unexpectedly found patients with severe illness secondary to SARS-CoV-2 infection to have monocytes with a blunted interferon response and dysregulation of antigen presentation as well as defective humoral and lymphocyte cytotoxic activity that could be contributing to the delayed viral clearance found in severely ill COVID-19 patients. The implications of these results are that treatment of patients with ARDS from SARS-CoV-2 infection may need a more targeted approach instead of using broad immunosuppression.

## Results

### Mild, severe, and recovering patient groups

We identified and collected blood for buffy coat samples from hospitalized patients confirmed to have SARS-CoV-2 infection (Supplemental Table 1). There were no significant differences in age, white blood cell count, cell subsets, and hematocrit between the three groups, but the platelet count was decreased in the severe versus mild group (Fig. 1A & 1B). Using available clinical data, we compared the expression of various inflammatory markers in the blood between groups at the time of sample collection and identified a significantly higher level of c-reactive protein in severe patients reflecting the hyperinflammatory state relative to mild patients (Fig. 1C).

**Figure 1.**
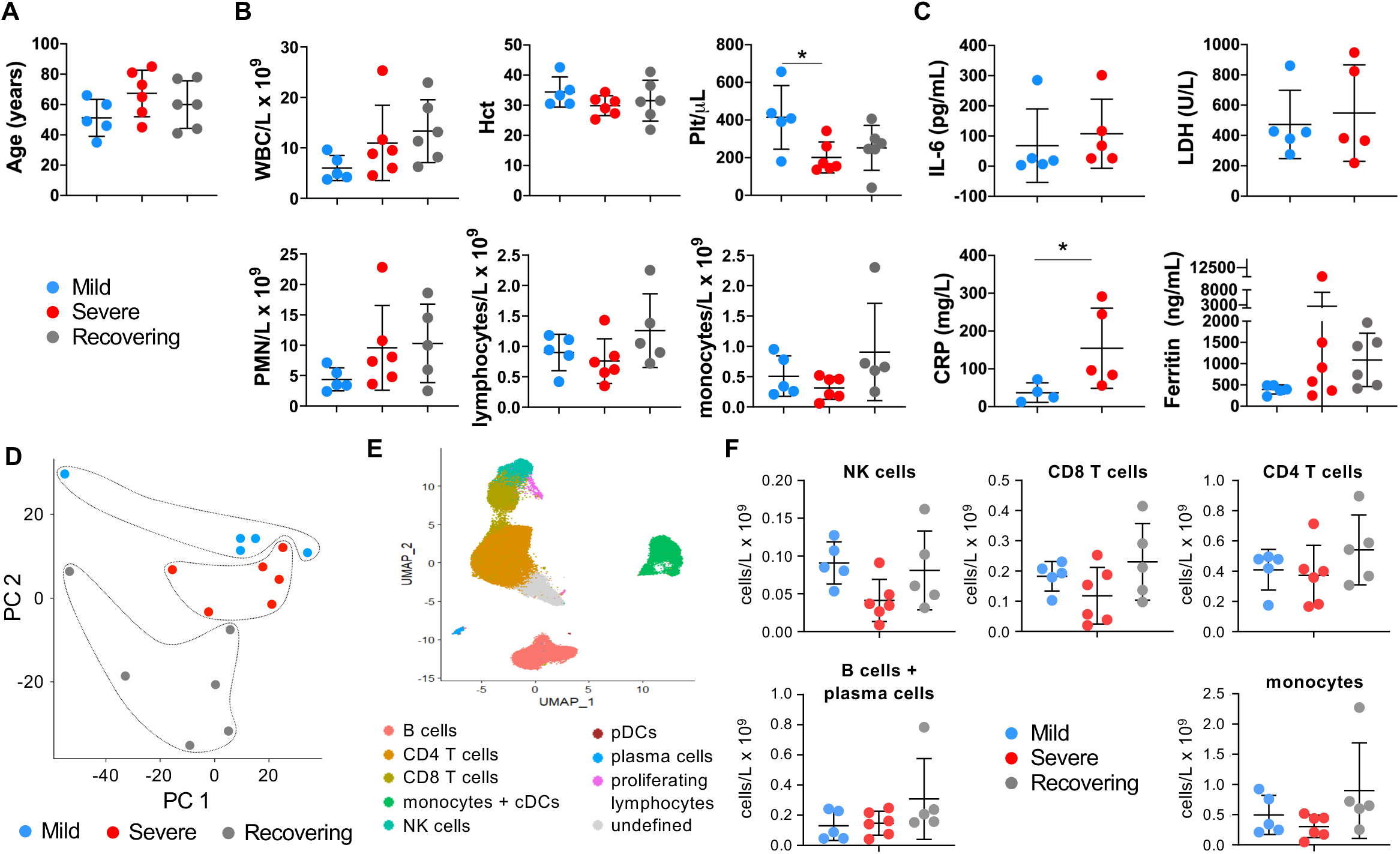
Evaluation of blood cells subsets in mild, severe (ARDS) and recovering (post-ARDS) COVID-19 patients. A) Age distribution of hospitalized COVID-19 patients requiring minimal respiratory support (mild, n=5), with ARDS (severe, n=6), and recovering from ARDS (recovering, n=6). B) Clinical CBC with differential cell count for COVID-19 patients. One-way ANOVA with Tukey’s multiple comparisons were used to test significance. C) Inflammatory markers in patient peripheral blood samples at admission (IL-6, LDH and CRP were not available for the recovering group). Mann-Whitney was used for IL-6, LDH, and CRP; and Kruskal-Wallis test with Dunn’s multiple comparison was used for ferritin. D-E) Peripheral blood leukocytes were assessed by scRNAseq. Principal component analysis (PCA) demonstrates clustering by disease severity (D). UMAP visualization reveals the major immune cell subsets (E and Supplemental Figure 1) (F) Cell counts of each major cell type. Proportions of leukocyte subsets detected by scRNAseq and clinical CBC from (B) were used to determine the numbers of specific cell types.

To identify functional differences in the peripheral blood leukocytes that may determine why some patients have milder disease courses whereas others develop ARDS, we used scRNAseq to survey the transcriptome of immune cells in mild, severe, and recovering patients. A principal component analysis (PCA) of the scRNAseq data from each sample demonstrated clustering by disease severity (Fig. 1D). The data was then visualized by Uniform Manifold Approximation and Projection (UMAP), and the predominant cellular populations in the peripheral blood were identified using gene expression of canonical markers (Fig. 1E and Supplemental Fig. 1). Clinical counts of lymphocytes and monocytes were used to estimate numbers of NK, CD4 T, CD8 T, B and plasma cells (all considered lymphocytes) and monocytes identified by scRNAseq (Fig. 1B and 1F). Consistent with their infrequency among circulating immune cells, insufficient conventional and plasmacytoid dendritic cells (cDCs and pDCs) were captured for analysis. Neutrophils were not identified because they, unlike other peripheral blood subsets, do not tolerate the cell freezing process ^16^

### Cytotoxic lymphocytes have a higher interferon response in severe patients but deficiency in cell killing

NK cells are an important arm of the innate lymphocytic antiviral response ^30^. We compared the mild versus severe group to identify differences in NK cell function that may determine why some have a more benign course of disease compared to those that develop ARDS (Supplemental Data Fig. 2). No significant differences in cell numbers were detected (Fig. 2A), yet pathway analysis revealed differences in the NK cell transcriptome between groups. Biological processes that were highly represented by genes upregulated and downregulated in severe patients compared to mild patients were identified (Fig. 2B and Supplemental Table 2 & 3). Patients with severe disease have higher viral loads ^31^. Accordingly, biological processes such as ” response to type I interferon”, ” response to virus”, ” response to interferon-gamma”, and ” response to interferon-beta”, among several others characteristic of a higher viral response were significantly upregulated in the severe group compared to mild.

**Figure 2.**
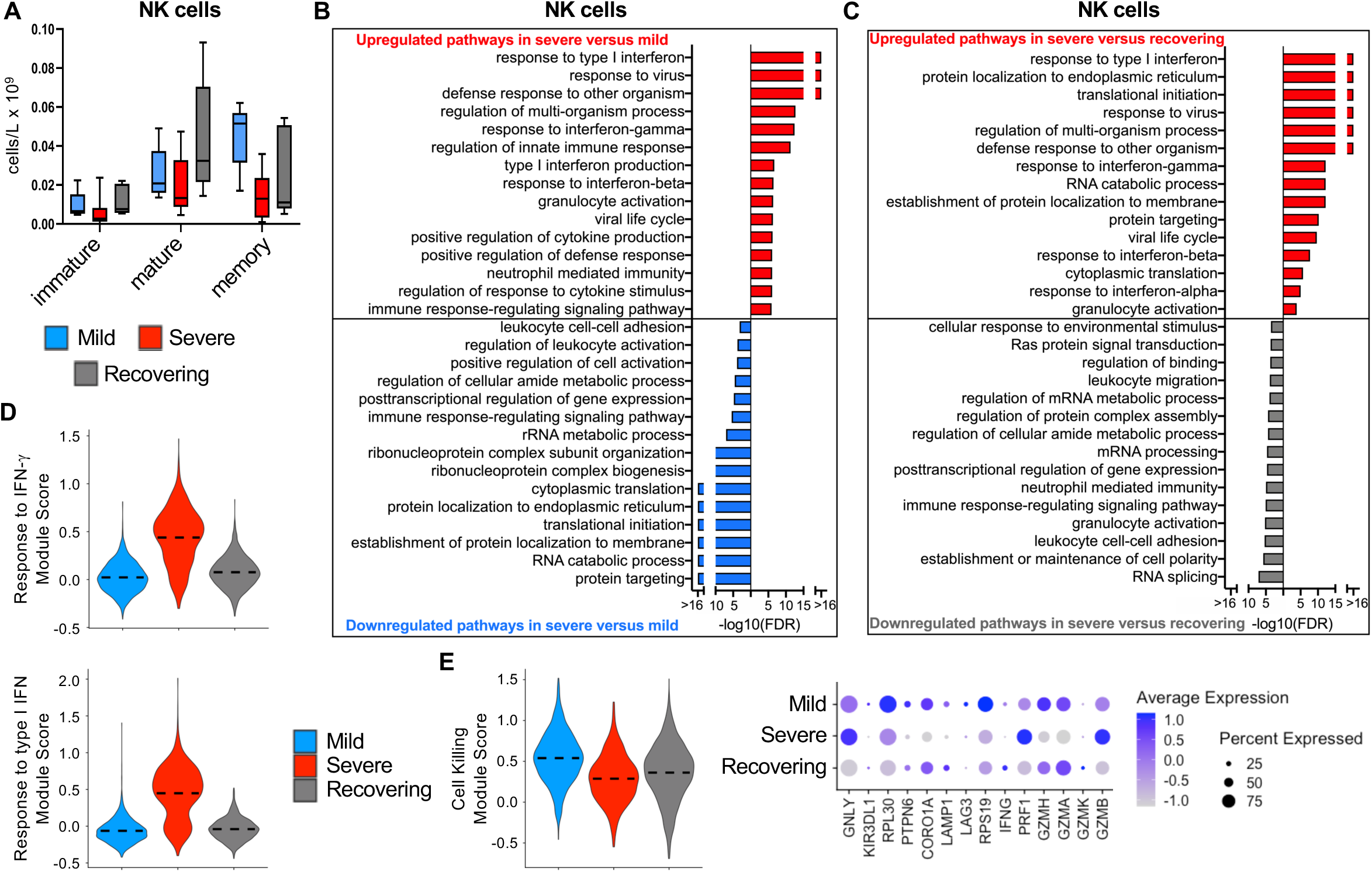
NK cells in severe patients have higher interferon signaling but dysfunctional cell killing. A) Subsets of NK cells were identified by their gene signatures and cell numbers were calculated using proportions of NK cell subsets and clinical lymphocyte count (see Figure 1B, Supplemental Figure 2A-D). B) Global transcriptome differences between severe and mild, and C) severe and recovering were evaluated in all NK cells by over representation analysis of up- and downregulated biological processes. D) Violin plots of response to IFN-*γ* and response to type I IFN modules of each cell from patient groups. E) Cell killing modules of each cell from patient groups and average expression of differentially expressed genes involved in cytotoxicity from patient groups. Kruskal-Wallis test was used to test significance in module scores, P<2.2^-16.^

Comparing critically ill patients with ARDS during their acute illness (i.e., severe group) to those that are recovering from severe disease could provide insight into pathways that drive the non-resolving inflammation in ARDS pathogenesis ^32^. Thus, we compared the biological processes that were affected by both up- and downregulated genes between severe and recovering groups (Fig. 2C and Supplemental Table 2 and 3). The recovering group had a resolution of the antiviral pathways, which would be expected from the waning viral load over time. When all three groups were evaluated together, a clear upregulation of the response to both type I and II interferons was found in the severe group in comparison to mild and recovering groups (Fig. 2D). Despite this response, which reflects higher viral recognition in NK cells in the severe group, genes associated with cytotoxic function were decreased in contrast to the mild and recovering group suggesting a dysfunctional effector antiviral response by NK cells in ARDS patients (Fig. 2E). Moreover, evaluating the data as individual patients (Supplemental Fig. 2) rather than all cells from the patient groups (Fig. 2D & 2E) revealed the same pattern of changes in interferon signaling and cytotoxic function.

We performed a similar evaluation of CD8 T lymphocytes to investigate the biological processes that differentiate the severity of disease and that are activated during the convalescent phase (Supplemental Fig. 3). There was large variability between patients, and no significant differences in cell numbers were detected (Fig. 3A). CD8 T cells had appropriate activation of interferon signaling that correlated with the disease severity (Fig. 3B – 3D, Supplemental Fig. 3E, and Supplemental Tables 4 & 5). We also found a gene signature indicating increased apoptosis in CD8 T cells, which is consistent with prior reports and could be driving the lymphopenia associated with COVID-19 illness (Fig. 3E) ^33^. Interestingly, CD8 T cells had a pattern of gene expression that suggested a deficiency in cytotoxic function in both severe and recovering groups in comparison to patients with milder disease (Fig. 3F). Although this pattern is expected in recovering patients that have resolving SARS-CoV-2 infection, the altered cytotoxic functional pathways in patients with ARDS mirrors the findings of NK cells (Fig. 2E).

**Figure 3.**
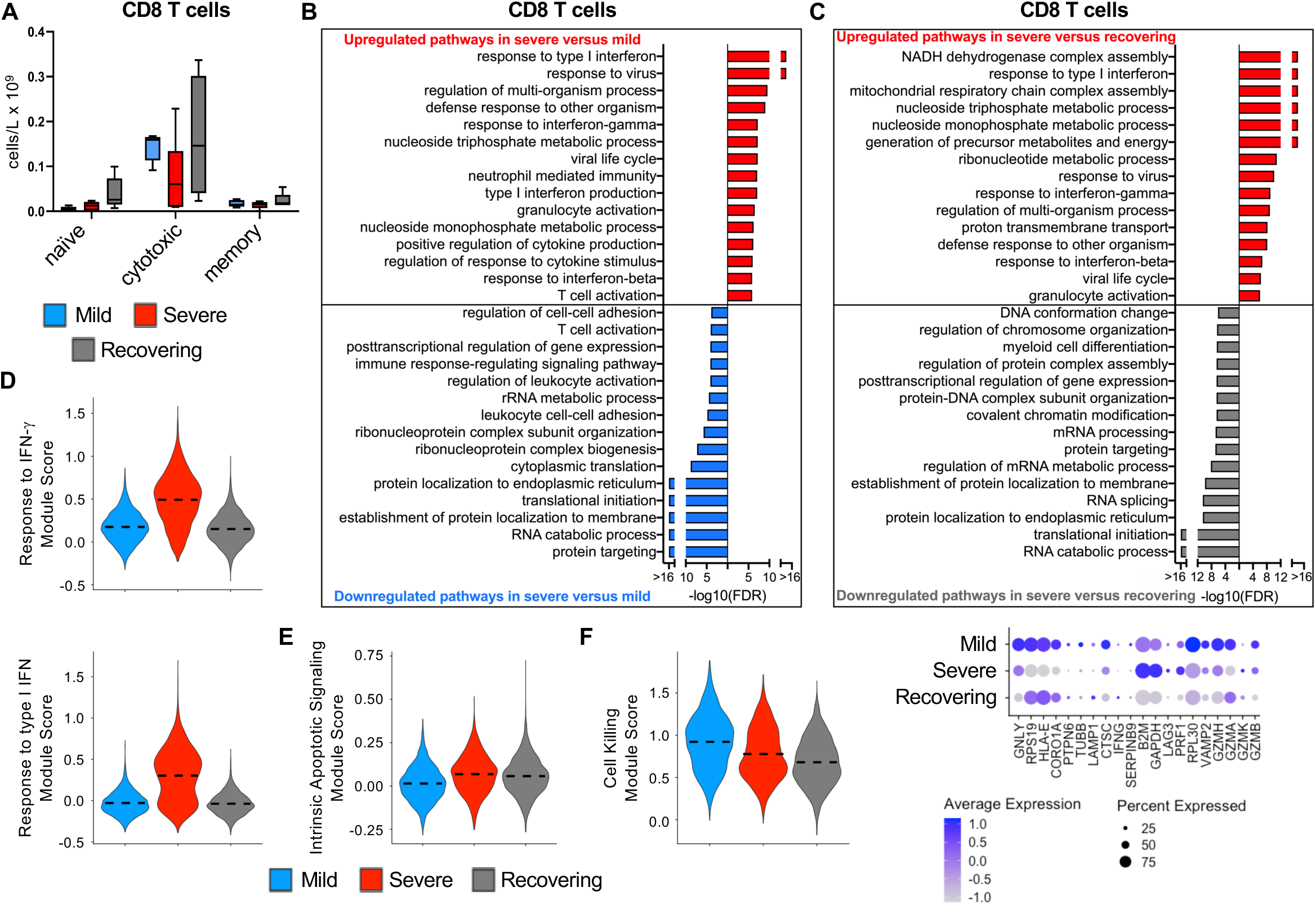
CD8 T cells in severe patients have higher interferon signaling, increased apoptotic gene expression, and dysfunctional cell killing. A) Subsets of CD8 T cells were identified by their gene signatures and cell numbers were calculated using proportions of CD8 T cell subsets and clinical lymphocyte count (see Figure 1B, Supplemental Figure 3A-D). B) Global transcriptome differences between severe and mild, and C) severe and recovering were evaluated in all CD8 T cells by over representation analysis of up- and downregulated biological processes. D-E) Violin plot of response to IFN-*γ* and response to type I IFN modules (D) and ntrinsic apoptotic signaling modules (E) of each cell from patient groups. F) Cell killing modules of each cell from patient groups and average expression of differentially expressed genes involved in cytotoxicity from patient groups. Kruskal-Wallis test was used to test significance, P<2.2^-16^.

Next, we evaluated CD4 T lymphocytes to determine up- and downregulated biological pathways in mild versus severe and severe versus recovering groups (Fig. 4A – 4C, Supplemental Fig. 4, and Supplemental Table 6 & 7). There were no significant differences in CD4 T cell subset numbers between groups (Fig. 4A), but consistent with NK and CD8 T lymphocytes, the severe group had a higher interferon response in CD4 T cells when compared to mild patients, and this was appropriately diminished in recovering patients (Fig. 4B-D). In addition to the antiviral response, several metabolic pathways were elevated in CD4 T cells in severe patients in comparison to other groups, and they also had more activation of apoptotic pathways (Fig. 4B, 4C, 4E, Supplemental Fig. 4E, and Supplemental Table 7).

**Figure 4.**
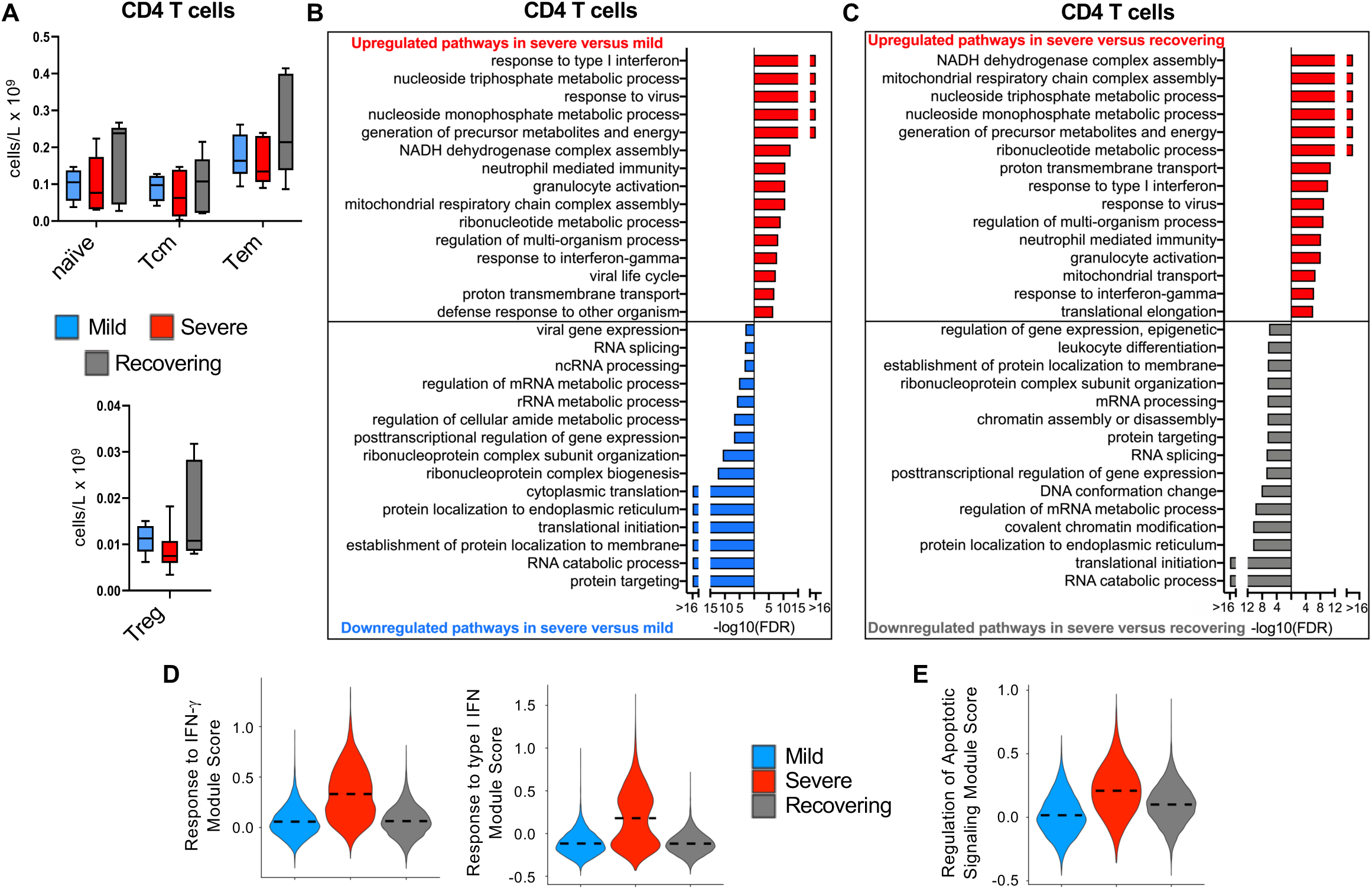
CD4 T cells in severe patients have higher interferon signaling, increased apoptotic gene expression, and are metabolically activated. A) Subsets of CD4 T cells were identified by their gene signatures and cell numbers were calculated using proportions of CD4 T cell subsets and clinical lymphocyte count (see Figure 1B, Supplemental Figure 4A-D). B) Global transcriptome differences between severe and mild, and C) severe and recovering were evaluated in CD4 T cells (all T cells expressing CD4) by over representation analysis of up- and downregulated pathways for biological processes. D-E) Violin plot of response to IFN-*γ* and response to type I IFN (D), and regulation of apoptotic signaling (E) modules of each cell from patient groups. Kruskal-Wallis test was used to test significance, P<2.2^-16^.

Altogether, these findings indicate that NK and T lymphocytes have an expected response to interferon signaling. Moreover, T lymphocytes are highly metabolic with increased activation of apoptotic pathways. However, both innate and adaptive cytotoxic lymphocytes have dysfunctional cytotoxic activity-associated pathways in severely ill COVID patients who have higher viral titers and delayed clearance of SARS-CoV-2 ^33^.

### B cells have dysregulated activation in severe patients

B lymphocytes also have an important role in the antiviral response ^34^. We identified B cells as well as their subsets to determine the differentially expressed genes between patient groups (Supplemental Fig. 5). Numbers of B cell subsets were not consistently different across groups (Fig. 5A). Similar to NK and T lymphocytes, the B lymphocytes also had higher interferon-mediated responses and activation of apoptotic signals in severe patients compared to both the mild and recovering groups (Fig. 5B – 5E, Supplemental Fig. 5E, and Supplemental Table 8 & 9). Moreover, B cell activation was upregulated in the recovering group in comparison to severe patients, which is the expected response during the convalescent phase of COVID infection (Fig. 5F). In contrast, B cells were less activated in the severe group when compared to mild patients indicating a delayed humoral response that could be contributing to the severity of disease (Fig. 5B & 5F).

**Figure 5.**
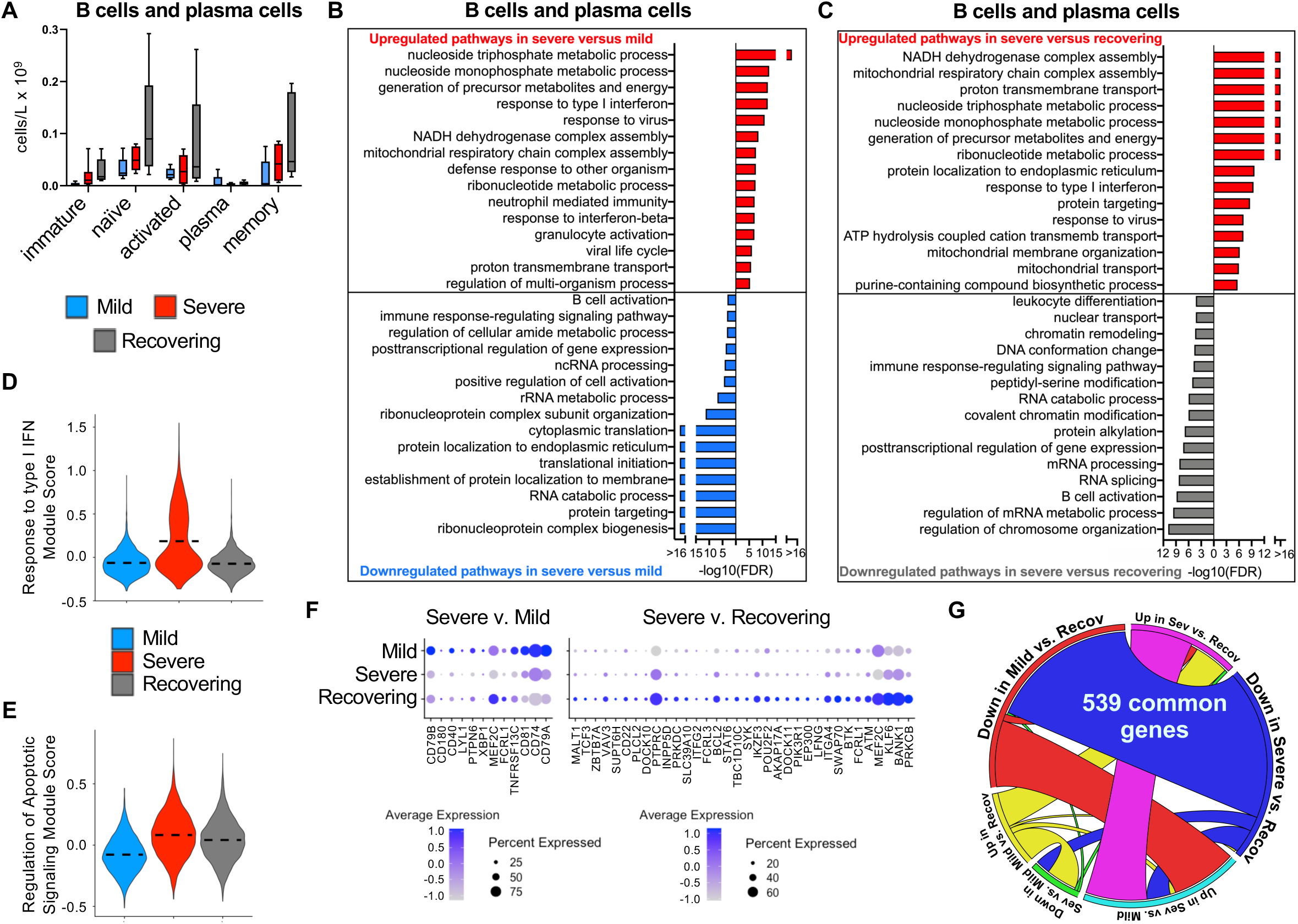
B cells in severe patients have higher interferon signaling, increased apoptotic gene expression, and dysfunctional activation signals. A) Subsets of B cells were identified by their gene signatures and cell numbers were calculated using proportions of B cell subsets and clinical lymphocyte count (see Figure 1B, Supplemental Figure 5A-D). B) Global transcriptome differences between severe and mild, and C) severe and recovering were evaluated in B and plasma by over representation analysis of up- and downregulated pathways for biological processes. D-E) Violin plot of response to type I IFN (D) and regulation of apoptotic signaling (E) modules of each cell from patient groups. Kruskal-Wallis test was used to test significance, P<2.2^-16^. F) Average expression of differentially expressed genes in the B cell activation pathway between mild and severe, and severe and recovering groups. G) Cord diagram showing the overlap of up and down regulated genes in the B cell population across COVID-19 groups.

In order to understand the pathways that are activated in B cells during the resolution from SARS-CoV-2 infection, we determined the common genes that changed in the comparison of mild versus recovering and severe versus recovering and identified 539 genes that were upregulated in the recovering group in both comparisons (Fig. 5G). The top canonical pathways enriched among these genes based on the Ingenuity Pathway Analysis (IPA) were in PI3K, IL-3, and IL-4 signaling, which are important signals required for B cell memory and the humoral response (Supplemental Fig. 6 – 8). We performed a causal network analysis to understand the upstream signals that regulated the differential upregulation of genes in the recovering group and found that SYK, which is a critical signal in B cell differentiation, maturation, and signal transduction by the B cell receptor, was the top upstream regulator candidate among all of the differentially expressed genes (Supplemental Fig. 9) ^35^.

### Monocytes are hyporesponsive in severe patients during SARS-CoV-2 infection

Emerging evidence demonstrates disruptions to the myeloid compartment in COVID-19 patients ^16^. We evaluated the monocyte subsets and found no differences in cell numbers (Fig. 6A). As expected, classical monocytes comprised the majority of the cells (Fig. 6B, Supplemental Fig. 10, Supplemental Table 10 & 11). Classical monocytes had a gene signature consistent with a decreased activation state, impaired phagocytosis, and altered differentiation in the severe compared to mild group (Fig. 6B & 6E). This functional deficiency in classical monocytes appears to recover during the resolution of ARDS suggesting that improvement in monocyte function could facilitate the resolution of inflammation in patients that develop ARDS (Fig. 6C & 6E). Moreover, monocytes in the severe groups had a dysfunction in antigen processing and presentation, including lower expression of MHC I and II (Fig. 6B, 6F, 6G). Additionally, in contrast to the lymphocytes, monocytes in severely ill patients had a deficient response to interferon signaling in comparison to mild and recovering groups that could at least partially explain the hypofunctional state that we identified in this cell compartment (Fig. 6D and Supplemental Fig. 10).

**Figure 6.**
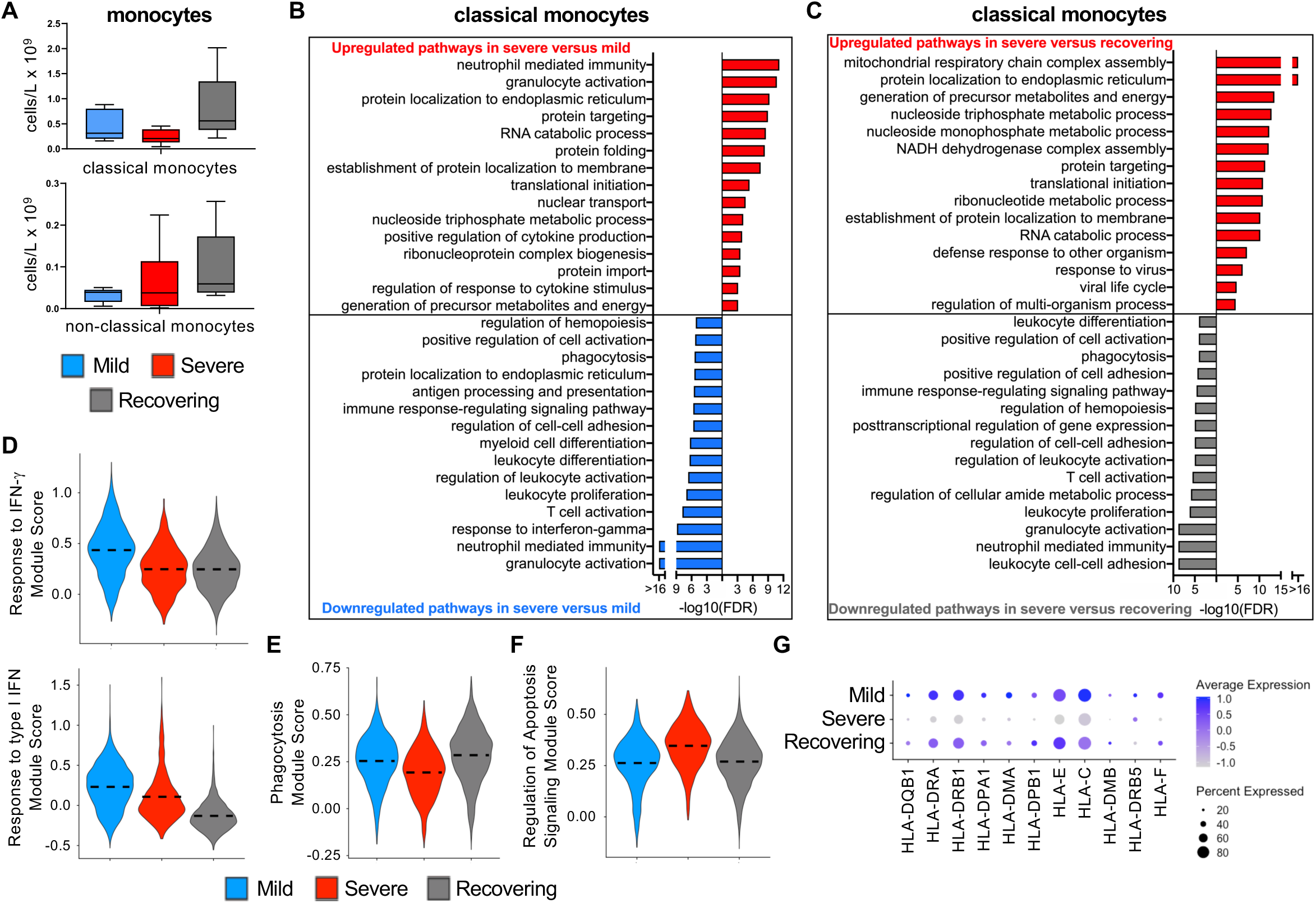
Classical monocytes in severe patients have impaired interferon signaling, phagocytosis, and antigen presentation. A) Subsets of monocytes were identified by their gene signatures and cell numbers were calculated using proportions of monocyte subsets and clinical monocyte count (see Figure 1B, Supplemental Figure 10A-E). Classical monocytes were analyzed further. B) Global transcriptome differences between severe and mild, and C) severe and recovering were evaluated in classical monocytes (CD14+CD16-) by over representation analysis of up- and downregulated pathways for biological processes. D-F) Violin plots of response to IFN-*γ* and response to type I IFN (D), phagocytosis (E), and regulation of apoptotic signaling (F) modules of each cell from patient groups. Kruskal-Wallis test was used to test significance, P<2.2^-16^. G) Average expression of differentially expressed HLA genes by classical monocytes from patient groups.

### Canonical pathways altered in various immune populations

To further evaluate the potential mechanisms that were driving the selective immunodeficiencies identified within various cell compartments, we performed a canonical pathway analysis with IPA (Supplemental Tables 12). In contrast to standard GO analysis where activation or inhibition of a process is inferred from up- and downregulation of its member genes respectively, canonical pathway activation incorporates prior knowledge of the overall consequences of up- and down-regulation of individual members of a process in activating or suppressing that pathway (e.g., downregulation of an inhibitor can lead to the activation of a process). In this analysis, we identified CD4 and CD8 T cells as having significant activation of the T cell exhaustion pathway in the severe versus mild group (Supplemental Table 12), which is consistent with other descriptions ^16^.

Next, we explored patterns of altered signaling pathway changes across all cell compartments (Fig. 7). Oxidative phosphorylation pathways were uniformly elevated in the severe group when compared to the mild and recovering groups likely reflecting the increased metabolic state with more severe infection. In contrast, sirtuin signaling pathways were suppressed in severe conditions across all cell compartments. Sirtuins are known mediators of antiviral defense ^36, 37^. As with our GO evaluation of biological processes, interferon signaling was decreased in monocytes when comparing severe to mild groups. Similarly, respiratory viral signaling pathways were also uniquely suppressed only within monocytes when comparing severe to mild groups. Notably, PD-1/PD-L1 signaling, an immunologic checkpoint inhibitory pathway, was higher in monocytes within the severe group which could be contributing to the suppressed response of this cellular compartment. Finally, eIF2 signaling was the most significantly downregulated pathway in all cells (except monocytes) when comparing severe to mild groups (Supplemental Fig. 11) whereas mTOR signaling was depressed in the severe versus recovering groups.

**Figure 7.**
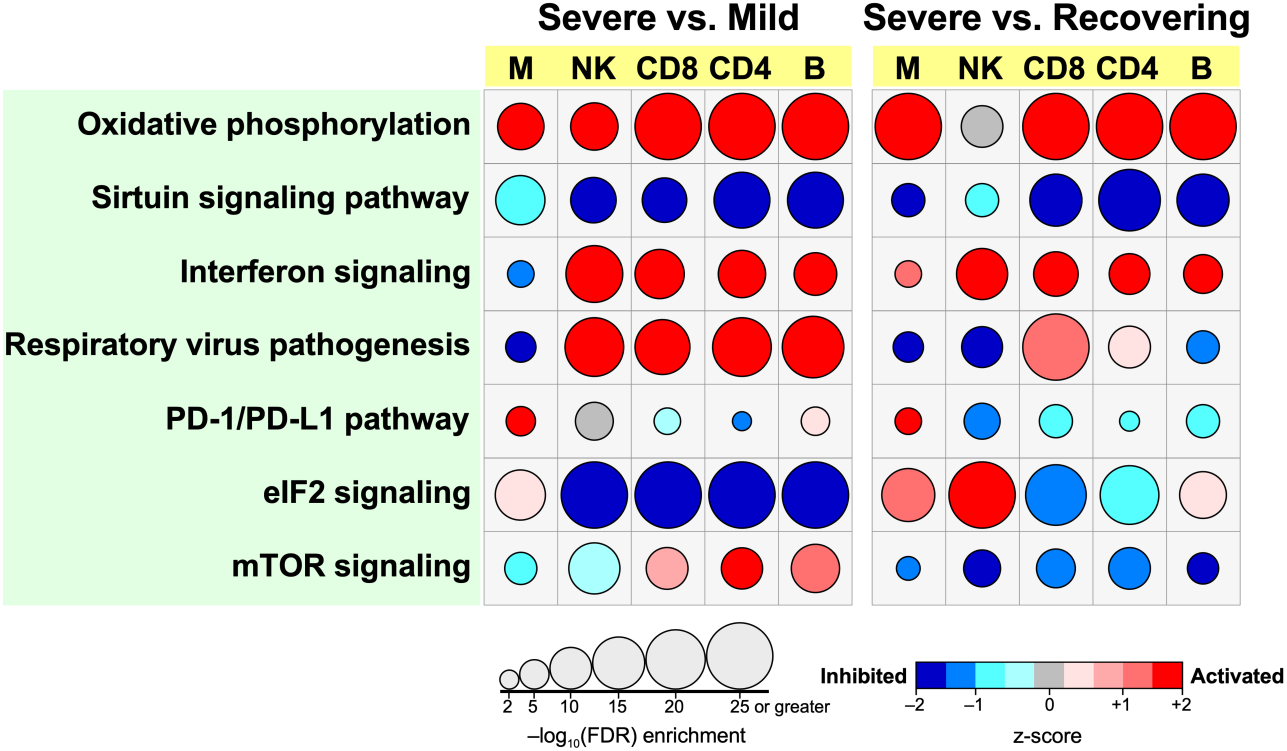
Canonical pathway analysis demonstrates defective signaling pathways across all immune cells in severe groups. IPA analysis of significantly enriched canonical pathways that were activated or inhibited between severe vs mild, and severe vs recovering groups in all cell types. FDR<0.01 was used to determine significance of enriched pathways, and z-score was used to determine pathways were activated or inhibited in the severe condition.

## Discussion

ARDS is driven by a dysregulated inflammatory response that has been largely described as a ” cytokine storm” associated with severely ill patients infected with SARS-CoV-2. A unique feature of the lung injury that perpetuates ARDS is a non-resolving inflammation even after the inciting factor, in this case SARS-CoV-2 infection, has resolved ^32^. In the recent months, a growing literature has developed a better understanding of how immune cell dysfunction contributes to the inflammatory response in COVID-19 patients ^9^. Several groups have adopted scRNAseq as an approach to functionally interrogate distinct immune compartments ^15, 16, 17, 18^. Our data are largely consistent with those findings, but our study further delineates the profound immune dysregulation in COVID-19 patients with severe illness compared to those with milder symptoms and those recovering from ARDS. Because our study obtained a sufficient number of samples within each group at more uniform disease timepoints, we were able to overcome heterogeneity in the data analysis from smaller sample sizes ^16^. By selecting patients groups that all had SARS-CoV-2 infection as opposed to comparing to healthy controls, we were also able to deeply interrogate immune cell dysfunction that drives the development of ARDS (i.e., mild versus severe) arising specifically from SARS-CoV-2 infection as well as understanding pathways that are activated during the resolution of inflammation (i.e., severe versus recovering). Using scRNAseq, we evaluated PBMCs and found that although most immune cellular compartments had an expected hyperinflammatory response in severe patients, presumably from the higher viral load and overall higher inflammatory state, several key pathways were dysfunctional in severe patients, which could be contributing to their inability to control the viral infection. Indeed, PBMCs from the severe group had deficiencies in functions necessary to clear virus: cytotoxic killing in NK and T cells, B cell activation, impaired antigen presentation by monocytes. Together, these data suggest functional deficiencies in innate and adaptive immune responses in SARS-CoV-2 infections that could be contributing to the severity of disease.

One of the first immunologic features identified in COVID-19 patients was decreased but hyperactive lymphocytes ^38, 39^. Our data suggest that both CD4 and CD8 T lymphocytes have a normal response to the viral pathogen with activation of antiviral pathways and a resultant hypermetabolic state. Notably, we found an increase in mitochondrial respiratory chain activity in the severe group, which could reflect a MAVS response to the SARS-CoV-2 infection ^40, 41^. It has also been suggested that lymphocytopenia results from increased apoptosis, which is consistent with our findings ^20, 42^. Additionally, our analysis indicates that CD8 T lymphocytes have a deficiency in cell killing that could be contributing to the pathobiology of COVID-19.

Similarly, NK cells had a transcriptomic signature that indicated patients with severe disease have defective effector killing function of virally infected cells. Clinical data on COVID-19 patients reported high viral load and impaired clearance in severe disease compared to mild cases ^43^. Studies in COVID-19 patients have found that SARS-CoV-2 persisted longer in respiratory samples of severe patients in comparison to those with milder symptoms, and this delayed viral clearance coincides with more lung damage from the infection ^31, 44^. Altogether, these data demonstrate a functional defect in both the innate and adaptive cytotoxic lymphocytic responses in COVID-19 patients with ARDS that could be contributing to the severity of disease.

Successful viral clearance relies on adaptive T cell and B cell immunity. Whereas cytotoxic T lymphocytes induce apoptosis of virus-infected cells, CD4 T cells activate B cells to differentiate into antibody-producing plasma cells and memory B cells ^45, 46^. Delayed IgM has been reported in severely ill COVID-19 patients suggesting a defect in B cell function, though it is unclear if this is due to direct defects in B cells or in CD4 T helper cells ^47^. Our findings suggest CD4 T lymphocytes have normal activity despite the decreased numbers described in COVID-19 patients. In contrast, pathways involved in B cell activation are downregulated in the severe group suggesting an inherent dysfunction in the B lymphocyte compartment that limits their activity. Moreover, SYK, which is essential for B cell development, was identified as the most significant upstream regulator that was downregulated in B cells within the severe group ^48, 49^. Although recovery from SARS-CoV-2 infection is not fully dependent on B cell function and antibody production ^50^, these immune cells likely still have an important role in controlling the severity of disease, and a dysfunctional humoral response may result in higher viral titers and delayed viral clearance that could contribute to the development of ARDS during SARS-CoV-2 infection.

Much attention has been devoted to lymphocyte dysfunction in COVID-19, particularly to the importance of cytotoxic T cells in overcoming viral infection ^27, 29, 39^. Yet the most dramatic difference we found in the severe group was a marked decline in monocyte function. We identified excessive activation of ER stress in monocytes with a concomitant increase in apoptosis, which could be augmenting death of this cellular compartment in the severe group. Peripheral blood monocytes also had a deficiency in their response to interferon signaling. Specifically, type I interferon activity is important during an appropriate antiviral immune response ^51^. However, a recent study demonstrated that severe COVID-19 patients have an apparent deficiency in type I interferon signaling ^17^. Our findings are congruous with this finding but also further reveal that the deficient type I interferon response emanates from the monocytic compartment in contrast to lymphocytes, which had an appropriate response. Monocytes in circulation from the severe group may be exhibiting immunoparalysis, as our results suggest they were not responsive to interferon signaling ^9^. Sudden loss of monocytes and their MHC expression is an indicator of severe acute illness in other conditions characterized by ” cytokine storm” such as sepsis and community acquired infections ^52, 53^. We also found monocytes to have decreased expression of class I and II MHC genes in the severe group consistent with other descriptions ^9, 16^, which may be related to reduced responsiveness to both type I IFN and IFN-*γ*. A concomitant augmentation of PD-1/PD-L1 signaling was found in monocytes, which could lead to the select immunosuppression in this immune population. Additionally, we postulate that suppressed protein translation by an augmented unfolded protein response in monocytes may be further contributing to the immunoparalysis phenotype identified in the Consequently, altered antigen presentation by monocytes can also result in the observed defect in T cell function in the severe group resulting in inhibition of the downstream adaptive immunity necessary for viral clearance and resolution of systemic inflammation ^54^.

Several cytokines have been found to be dysregulated in COVID-19 patients ^42^. Type I interferons are crucial for a successful antiviral immune response, and subverting the early type I interferon response contributes to pathogenesis in MERS-CoV, SARS-CoV-1, and SARS-CoV-2 infections ^12, 17, 18, 55^. Clinical trials with type I interferon therapy are ongoing, however, this treatment may be harmful if patients are already upregulating interferon stimulated genes. In fact, our pathway analysis suggests adaptive immune cells have a normal interferon response, but monocytes have a selective dysregulation with a depressed interferon response in severe COVID-19 patients. Furthermore, our findings suggest that monocytes in severe patients have an inherent defect in their ability to respond to interferon signaling, and this dysfunction may not be ameliorated by simply giving type I interferons to patients at high risk for developing ARDS from SARS-CoV-2 infection ^51^. IL-6 is another prominent cytokine that is elevated in the serum of COVID-19 patients and has been tied to the pathogenesis of lung injury. As such, IL-6 blockade (e.g., tocilizumab) has been used in the treatment of COVID-19 patients, and retrospective studies support this as a promising treatment in more severely ill COVID-19 patients ^56, 57^. Interestingly, IL-6 suppresses MHC class II expression on monocytes ^9, 58^, and treatments such as tocilizumab may not be uniformly immunosuppressive and could be improving the monocyte dysfunction identified by our analysis.

SARS-CoV-2 infection of immune cells could be causing the dysregulated response in PBMCs. However, immune cells do not express ACE2, the receptor required for viral entry, suggesting an inability for viral infection ^59, 60, 61, 62^. Moreover, we did not find any evidence of ACE2 expression or any viral RNA in our scRNAseq of PBMCs (unpublished). The alternative explanation is that genetic, environmental, and age-related changes could confer a predisposition to a dysregulated response to pro-inflammatory signals. This concept has been previously demonstrated where certain individuals having attenuated while others have augmented inflammatory responses upon LPS stimulation of PBMCs ^63^. Aging, a risk for more severe COVID-19 illness, could be contributing to immunoparalysis ^5, 6, 64^. Indeed, sirtuin signaling was uniformly depressed in all immune cell compartments of the severe group irrespective of comparison to mild or recovering patients. Sirtuins are histone deacetylases that have diverse effects in antiviral defense and controlling longevity, inflammation, and cellular senescence ^36, 37, 65^. In particular, sirtuin activity decreases with age, and immunosenescence could be driving some of the dysregulated immune response in severe patients ^64^. Obesity is another risk factor for development of severe COVID-19 disease ^5^, and immunometabolic alterations could be contributing to the dysfunctional immune response in COVID-19 patients. The immunomodulatory effects of obesity can alter both the innate and adaptive arms of the immune system and have profound effects in tissue inflammation ^66^. Autophagy has a prominent role in immunometabolism ^67^, and our findings demonstrate disrupted mTOR signaling in multiple cellular compartments of severe patients particularly when compared to recovering patients suggesting another possible link to the dysregulated immunologic findings in COVID-19 patients.

In summary, our study supports the concept that COVID-19, and especially severe cases that have progressed to ARDS, is characterized by multifaceted immune dysregulation that is not uniformly hyperinflammatory but more accurately described as a state of immune imbalance. Although immune cells have augmented inflammatory signatures in severe patients compared to the mild and recovering groups, they also have distinct patterns of immunoparalysis such as impaired cytotoxic cell killing, attenuated B cell activation, and dysregulated monocyte antigen presentation that could contribute to the severity of illness. Future research in immune therapies should consider a nuanced approach, particularly those involving targeted augmentation of pathways within specific immune compartments to limit the disease severity as well as promote resolution of the unrelenting inflammation in ARDS.

## Data Availability

All the sequencing data along with their associated metadata and accession code will be made available upon peer-reviewed publication.

## Acknowledgments

We thank the staff of the Biobank & Translational Research Core, the Flow Cytometry Core, and the Applied Genomics, Computation & Translational Core for their help in this project. This work is supported by funding by the Parker B. Francis Foundation Fellowship (CY), Plum Foundation Funding (PC), Erwin Rautenberg Foundation Fellowship (TP), UCLA CTSI KL2 (TP), and NIH grants T32HL134637 (SAB, JAP), R01AI134987 (HSG), AI137111 (SAG), and R01HL137076 (PC).

## Methods

### Ethics statement

This study was approved by Institutional Review Board at Cedars-Sinai Medical Center (CSMC; IRB# STUDY00000602). Informed consent was obtained from all enrolled patients according to CSMC BioBank’s Phase 2 Protocol (IRB# PRO00043021).

### Study design and sample collection

Patients admitted to CSMC and diagnosed with COVID-19 by RT-PCR of nasopharyngeal swabs were stratified into mild, severe, and recovery groups (n=6/group). One sample in the mild group was lost during processing. Mild disease was defined as having minimal oxygenation requirements (≤4 liters via nasal cannula) at the time of sample collection. Severe disease included critically ill patients admitted to the medical intensive care unit and requiring mechanical ventilation for acute respiratory distress syndrome (ARDS) during their hospitalization. Instead of sample collection based on days of symptoms, which is subjective and liable to variability, we chose to collect samples within 5 days of admission from mild and severe groups to maintain uniformity of timing for comparison between groups based on disease severity. Furthermore, we collected samples from a recovery group comprised of patients 18 to 25 days after admission who were recovering from ARDS secondary to SARS-CoV-2 infection. Venous blood was collected into EDTA coated tubes and centrifuged to separate plasma and buffy coat. Plasma was collected and frozen at -80C, and the buffy coat was collected into cryo-preservation media and frozen at -80C.

### Sample processing and single cell RNAseq

The frozen buffy coats were thawed and washed with PBS containing 10% FBS. The cells were stained with DAPI (3µM) for 5 min to assess viability. Live cells were sorted using a BD FACS Aria III (BD Biosciences, San Jose, CA, USA) in the CSMC Flow Cytometry Core. Totalseq-A human hashing antibodies (Biolegend) were used to label FACS sorted cells. Pairs of hashed samples from the same patient group were mixed together at a 1:1 ratio before methanol fixation using the 10X Genomics methanol fixation protocol ^68^. Single cells were captured using a 10X Chromium Controller (10X Genomics) and libraries were prepared according to the Single Cell 3’ Next GEM V3.1 Reagent Kits User Guide (10X Genomics). The barcoded sequencing libraries were quantified by quantitative PCR using the KAPA Library Quantification Kit (KAPA Biosystems, Wilmington, MA). Libraries were sequenced using a Novaseq 6000 (Illumina) with custom sequencing settings of 28bp and 91bp for read 1 and 2, respectively, to obtain a sequencing depth of ∼5×10^4^ reads per cell.

### Alignment, Demultiplexing, Quality Control and Batch Correction

CellRanger v3.0.0 software was used with the default settings for demultiplexing, aligning reads with STAR software to a custom human GRCh38 transcriptome reference downloaded from http://www.gencodegenes.org, containing all protein coding and long non-coding RNA genes based on human GENCODE version 33 annotation with SARS-Cov2 virus genome MT246667.1, https://www.ncbi.nlm.nih.gov/nuccore/MT246667.1. Cell hashing data were demultiplexed using CITE-seq-Count 1.4.3 ^69^.

Single cell analysis R package Seurat v3.1.5 was used for data analysis ^70^. For quality control and filtering out low quality cells, only cells expressing more than 200 genes (defined as genes detected in at least 3 cells) and fewer than 20% mitochondrial genes were selected. To minimize doublet contamination for each dataset, we removed high UMI cells by a fit model generated from suggested mulitplet rate over number of cells recovered as in the 10X Genomics user manual. Ambient RNA derived from lysed cells was removed using SoupX package ^71^. A total number of 69983 captured single cells (about 4000 cells per patient) passed quality control for further batch correction and unbiased clustering. Batch correction package Harmony with Seurat 3 wrapper was used for data integration ^72^. The batch correction was processed with PCA (Principal Component Analysis) using the 5000 most variable genes, and the first 20 independent components were used for downstream unbiased clustering with a resolution of 0.4. The UMAP (Uniform Manifold Approximation and Projection) method was used for visualization of unsupervised clustering. Cell cluster identities were determined using known gene markers of individual cell types. Differentially expressed genes between different clusters and groups were calculated using Mode-based Analysis of Single-cell transcriptomics (MAST) ^73^. Erythroid and Platelet clusters were removed from further analysis. Lymphocytes clusters without obvious subset markers were annotated as undefined lymphocytes.

### Immune cell counts

Lymphocyte and monocyte differential counts from clinical tests were used to estimate cell counts for each immune cell subset. Cells identified as NK cells, CD4 T cells, CD8 T cells, B and plasma cells, proliferating lymphocytes and undefined lymphocytes were collectively considered ” total lymphocytes”. Lymphocyte subset counts were estimated as: *subset cell count ÷ total lymphocyte count × lymphocyte differential count*. Classical and non-classical monocytes as well as cDCs and pDCs (which have similar morphology for differential counting) were collectively considered ” total monocytes”. Monocyte subset counts were estimated as: *subset cell count ÷ total monocyte count × monocyte differential count*. One sample in the recovering patient group lacked differential counts and was therefore excluded from the analysis of immune cell counts.

### Pathway analysis and module scores

To understand the biological processes that were enriched by the differentially expressed genes within each immune compartment, we sorted differentially expressed genes (adjusted *p* value < 0.01) into those that were upregulated or downregulated between two groups (i.e., mild versus severe and severe versus recovering) and did further analysis of gene lists separately. Using Webgestalt (Version 2019) ^74^, gene lists were entered to do an over-representation analysis using the gene ontology (GO) database to identify the biological processes that were affected by the differentially expressed genes between severity groups in each cell type.

In order to complement our GO analysis, we also performed canonical pathway analysis on all major immune cell types using Ingenuity Pathway Analysis (IPA). Enrichment analysis on differentially expressed genes (FDR < 0.01) for each cell type (Classical monocytes, NK, CD8 T cell, CD4 T cell, and B cell) across conditions (Mild, Severe, Recovering) was performed by leveraging over 700 manually curated pathways using Fisher’s exact test with Benjamini Hochberg adjusted *P*-values (FDR). Additionally, pathway activity analysis was performed to assess whether significantly enriched pathways (FDR < 0.01) were activated or inhibited based on IPA’s knowledgebase of expression or phosphorylation patterns of gene products in a given canonical pathway using a z-score statistic. While each cell type and condition elicited a distinct set of enriched and differentially activated/inhibited pathways, we focused on the most significant programs shared by immune cells under different clinical states.

Pathway module scores were determined using the union of differentially expressed genes in all groups returned from over-representation analysis for biological processes of interest. Pathway module scores were calculated using the AddModuleScore function of the Seurat package that calculated the average expression of each gene signature list and subtracted by the aggregated expression of control feature sets. All analyzed features are binned based on averaged expression, and the control features are randomly selected from each bin ^75^.

---

**Supplemental Figure 1.**
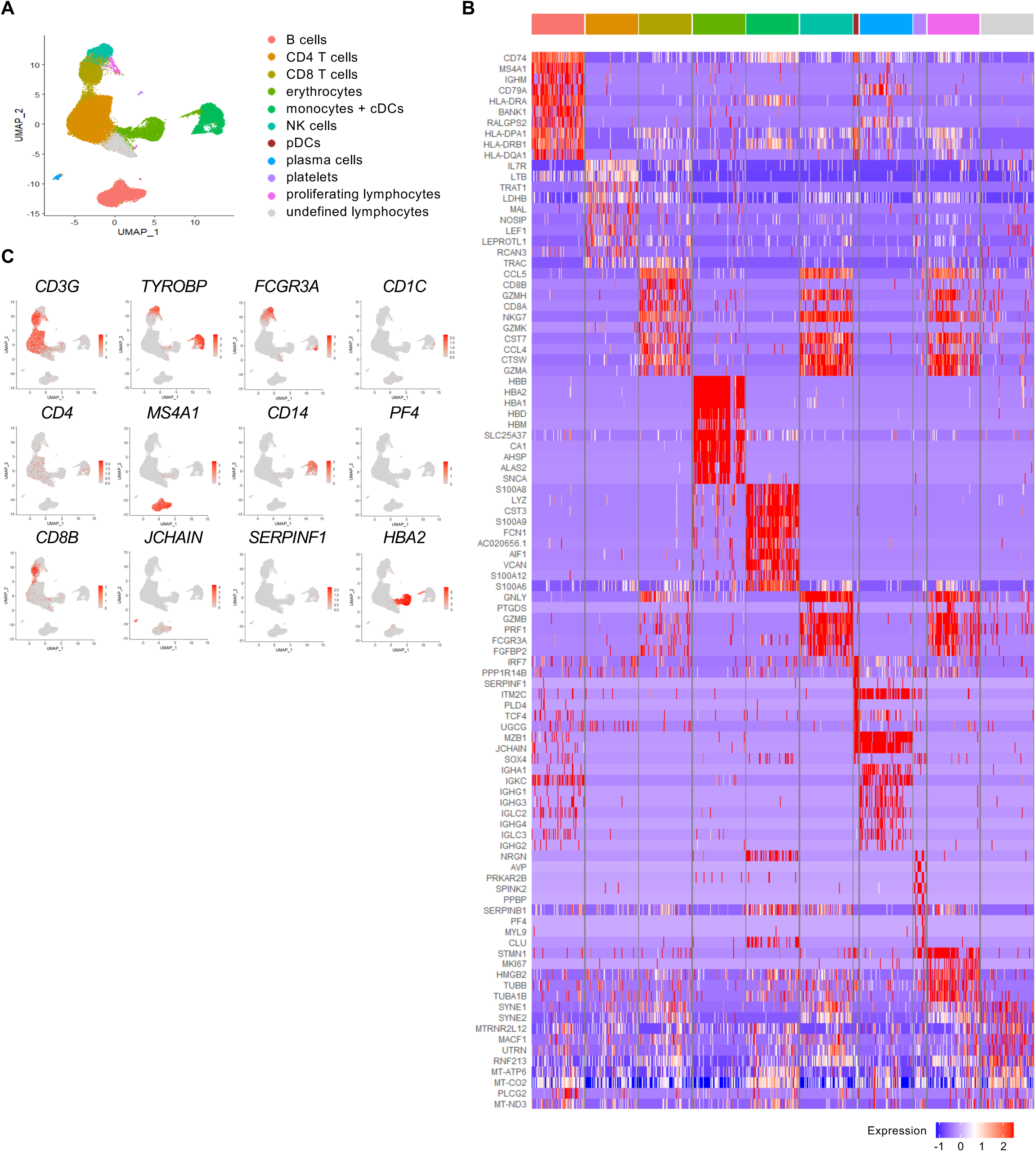
Identification of immune cell subsets. A) UMAP plot of all cells from all patients. B) Heatmap of genes differentially expressed by blood cells (downsampled). C) UMAP plots identify T cells (CD3G), NK cells (TYROBP, FCGR3A), B cells (MS4A1), plasma cells (JCHAIN), proliferating lymphocytes (MKI67 and CD3G or FCGR3A), monocytes (CD14 or FCGR3A), cDCs (CD1C), pDCs (SERPINF1), platelets (PF4) and erythrocytes (HBA2). Erythrocytes and platelets were excluded from further analysis.

**Supplemental Figure 2.**
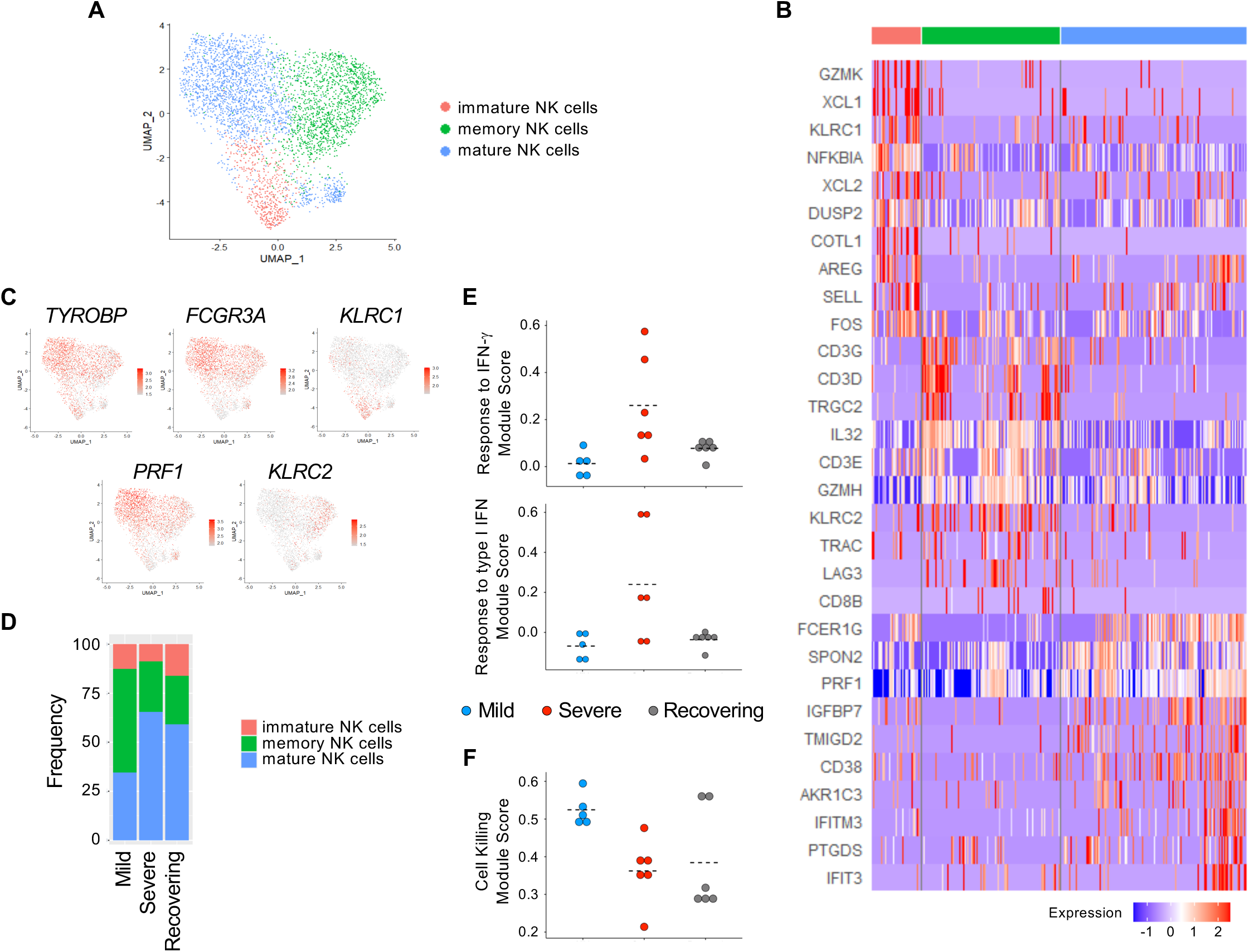
Identification of NK cell subsets. A) UMAP of NK cells from all patients. B) Heatmap of genes differentially expressed by NK cell subsets (downsampled). C) NK cell clusters were identified as immature (KLRC1), mature (FCGR3A, PRF1) and memory (FCGR3A, KLRC2) cells. D) Proportions of NK cell subsets in mild, severe and recovering patients. E-F) Mean module scores for individual patients.

**Supplemental Figure 3.**
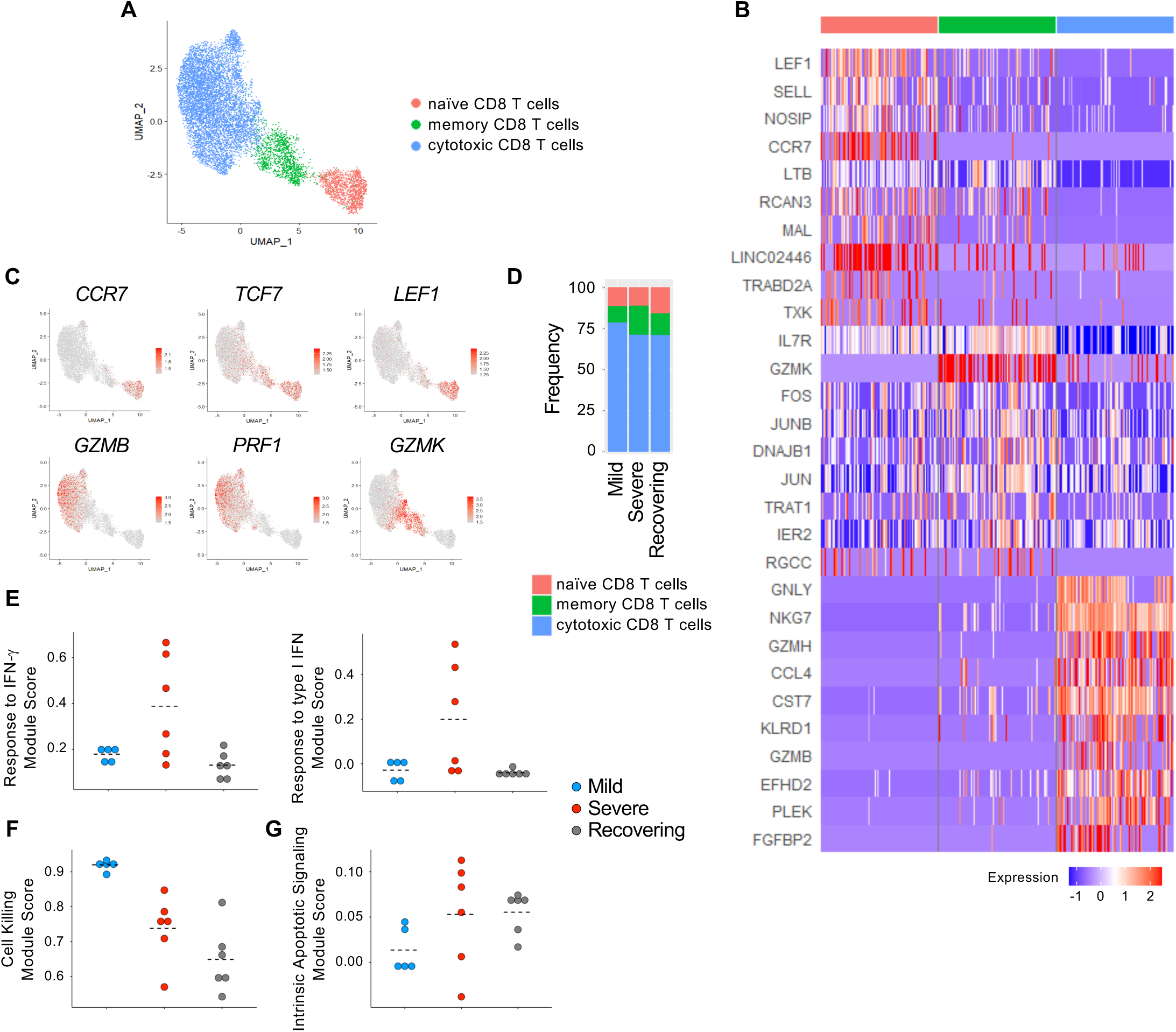
Identification of CD8 T cell subsets. A) UMAP of CD8 T cells from all patients. B) Heatmap of genes differentially expressed by CD8 T cell subsets (downsampled). C) CD8 T cell clusters were identified as naïve (CCR7, TCF7, LEF1), cytotoxic (GZMB, PRF1) and memory (GZMK) cells. D) Proportions of CD8 T cell subsets in mild, severe and recovering patients. E-G) Mean module scores for individual patients.

**Supplemental Figure 4.**
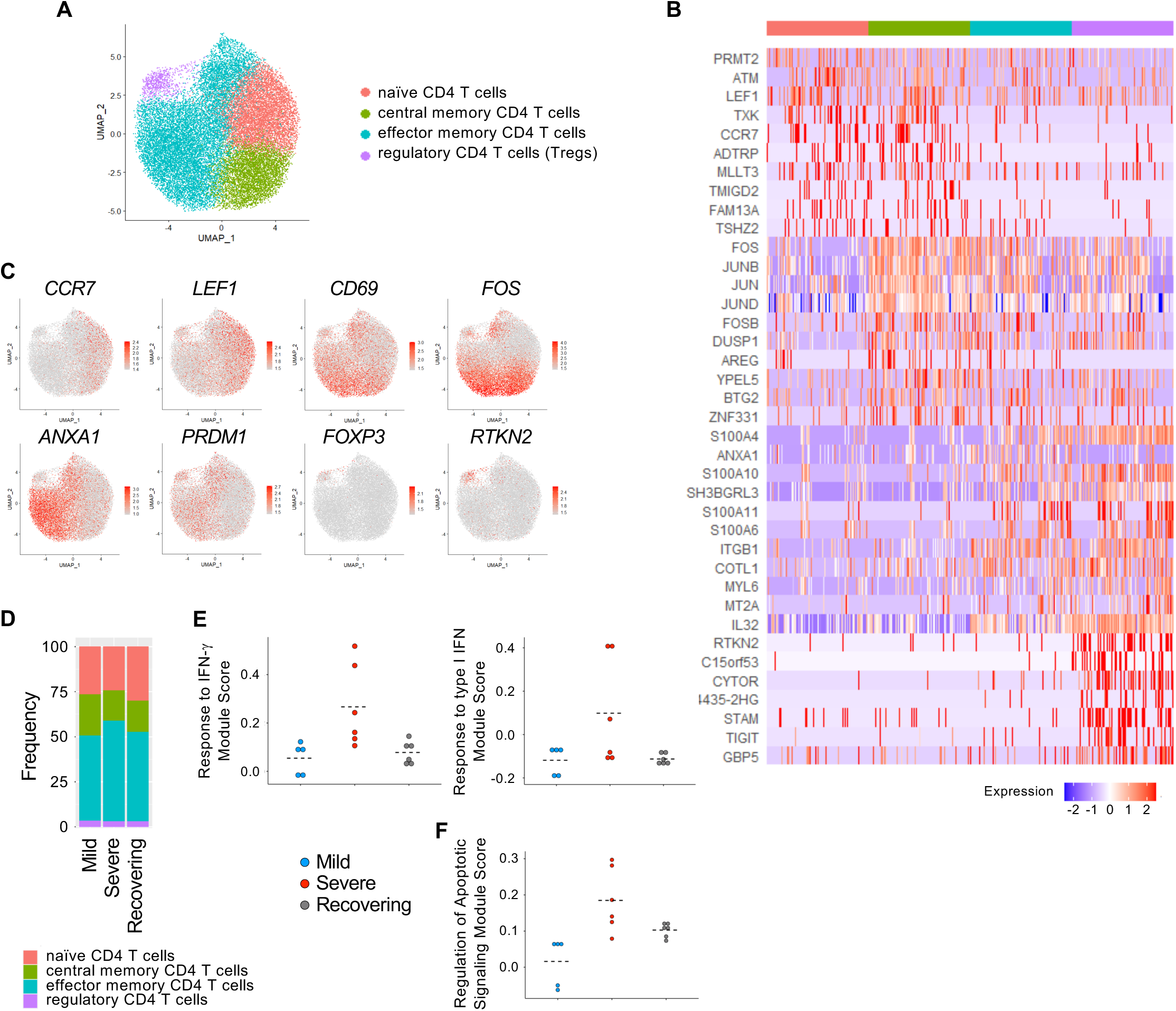
Identification of CD4 T cell subsets. A) UMAP of CD4 T cells from all patients. B) Heatmap of genes differentially expressed by CD4 T cell subsets (downsampled). C) CD4 T cell clusters were identified as naïve (CCR7, LEF1), central memory (CCR7, CD69), effector memory (ANXA1, PRDM1) cells and regulatory T cells (Tregs; FOXP3, RTKN2). D) Proportions of CD4 T cells subsets in mild, severe and recovering patients. E-F) Mean module scores for individual patients.

**Supplemental Figure 5.**
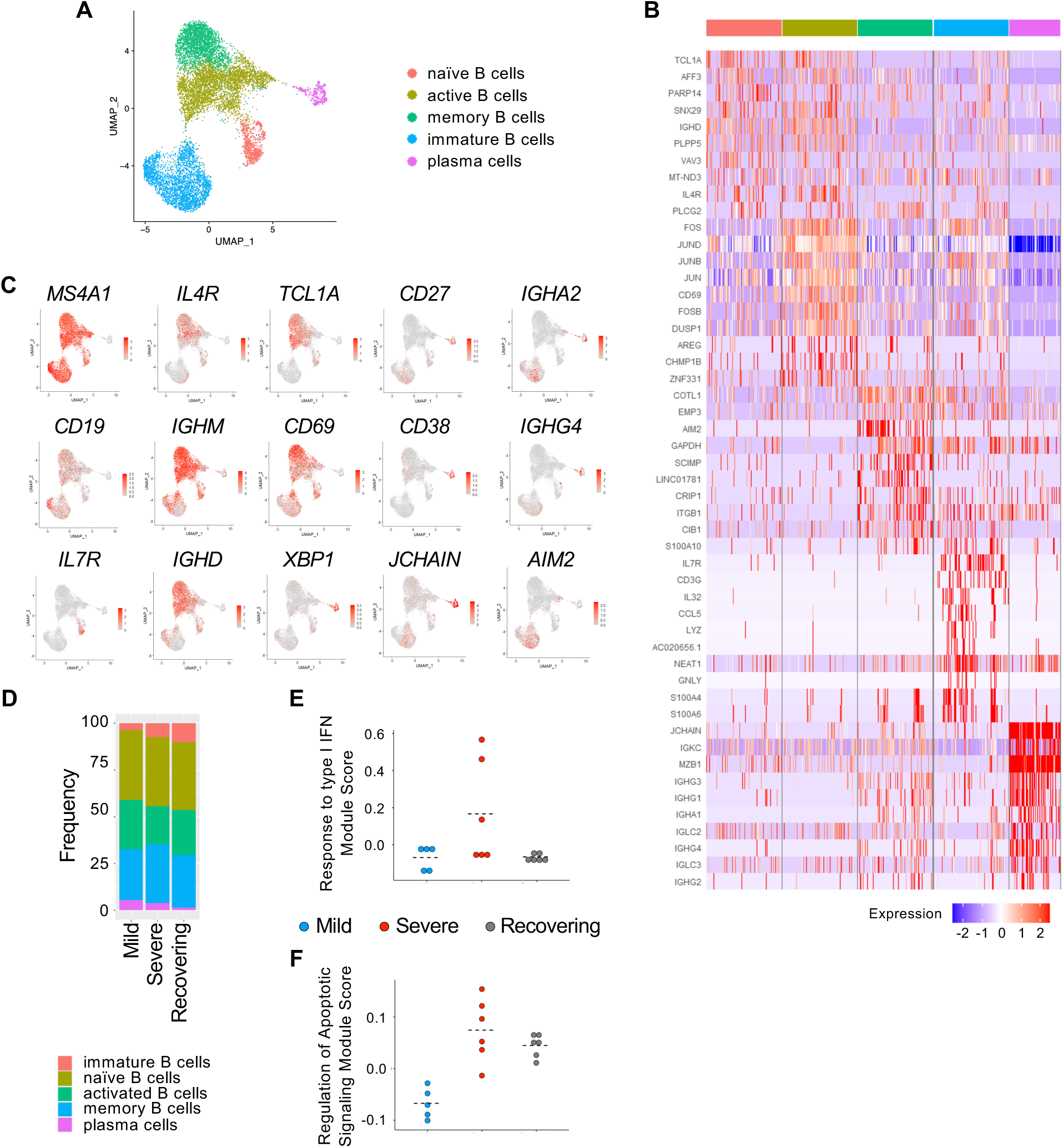
Identification of B and plasma cell subsets. A) UMAP of B and plasma cells from all patients. B) Heatmap of genes differentially expressed by B and plasma cell subsets (downsampled). C) B and plasma cell clusters were identified as immature B cells (IL7R), naïve B cells (IGHM, IGHD, IL4R, TCL1A), activated B cells (CD69), plasma cells (CD27, CD38, XBP1, JCHAIN) and memory B cells (AIM2). D) Proportions of B and plasma cells subsets in mild, severe and recovering patients. E-F) Mean module scores for individual patients.

**Supplemental Figure 6.**
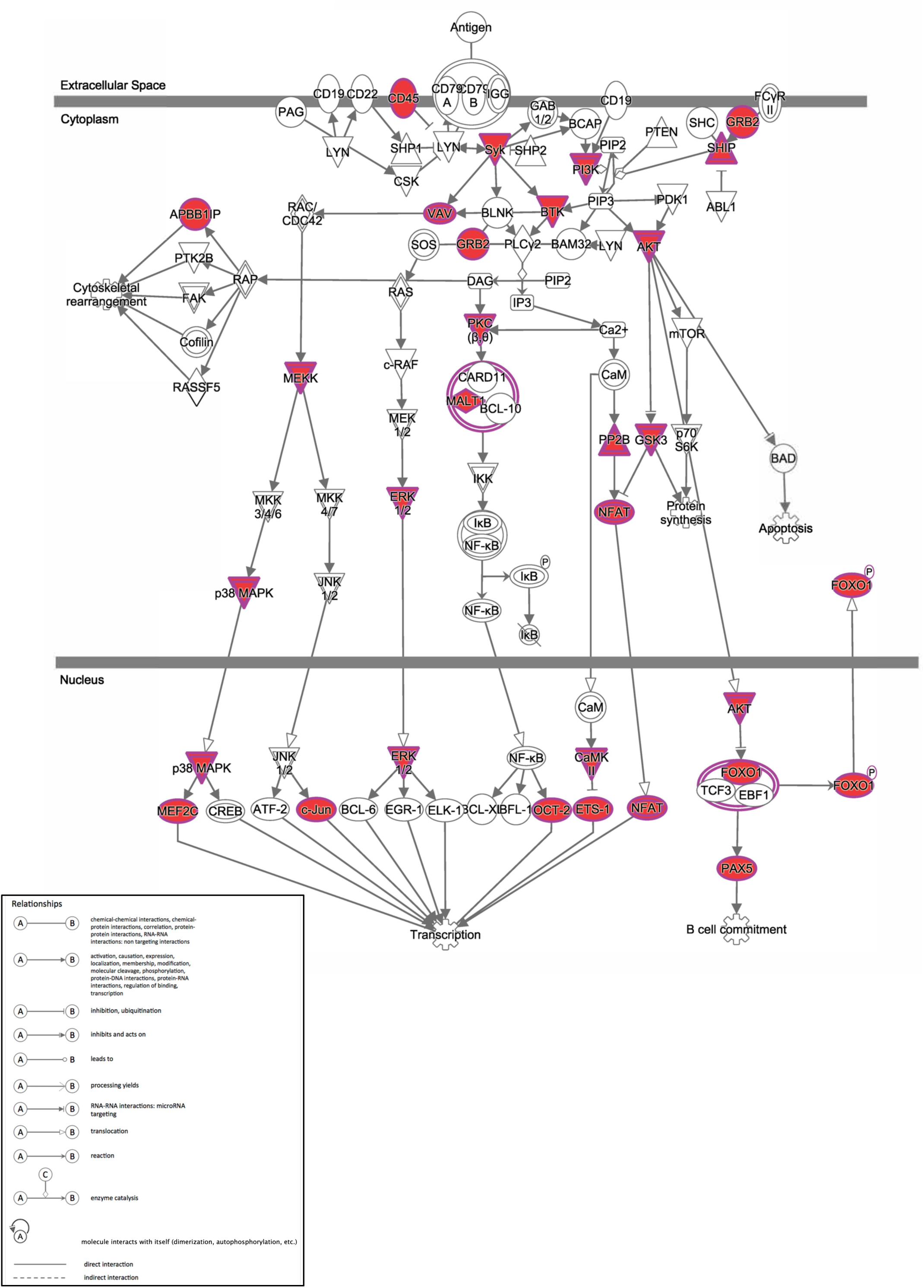
B cell receptor signaling is enriched in the recovering group. IPA canonical pathway analysis of genes significantly upregulated in B cells from recovering vs mild and severe COVID-19 patients. Analysis of the 539 DEGs upregulated (red) only in the recovering group shows increased signaling through the B cell receptor compared to mild and severe groups.

**Supplemental Figure 7.**
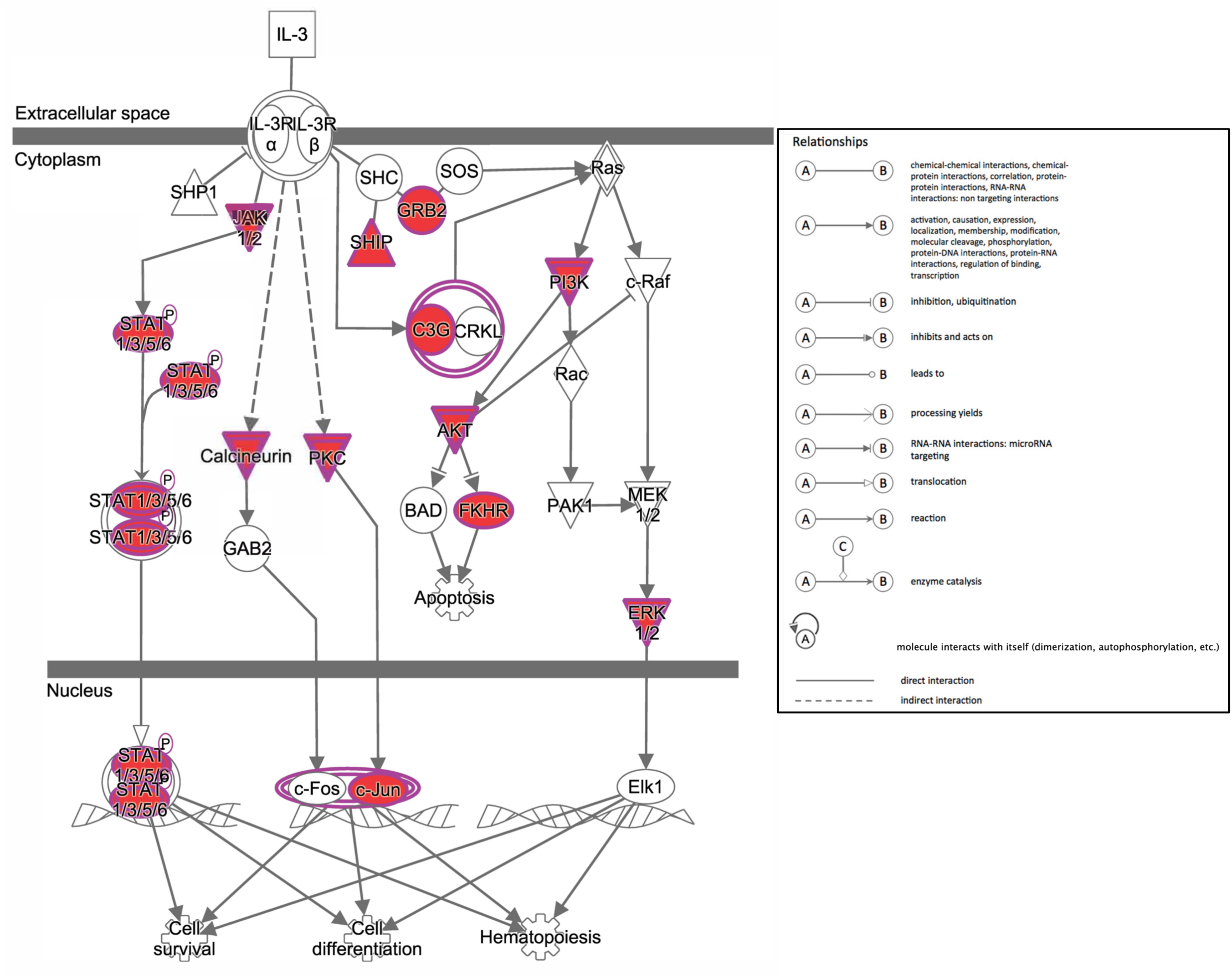
IL-3 signaling in B cells is enriched in recovering groups. IPA canonical pathway analysis of genes significantly upregulated in B cells from recovering vs mild and severe COVID-19 patients. Analysis of the 539 DEGs upregulated (red) only in the recovering group shows increased IL-3 signaling in recovering compared to mild and severe groups.

**Supplemental Figure 8.**
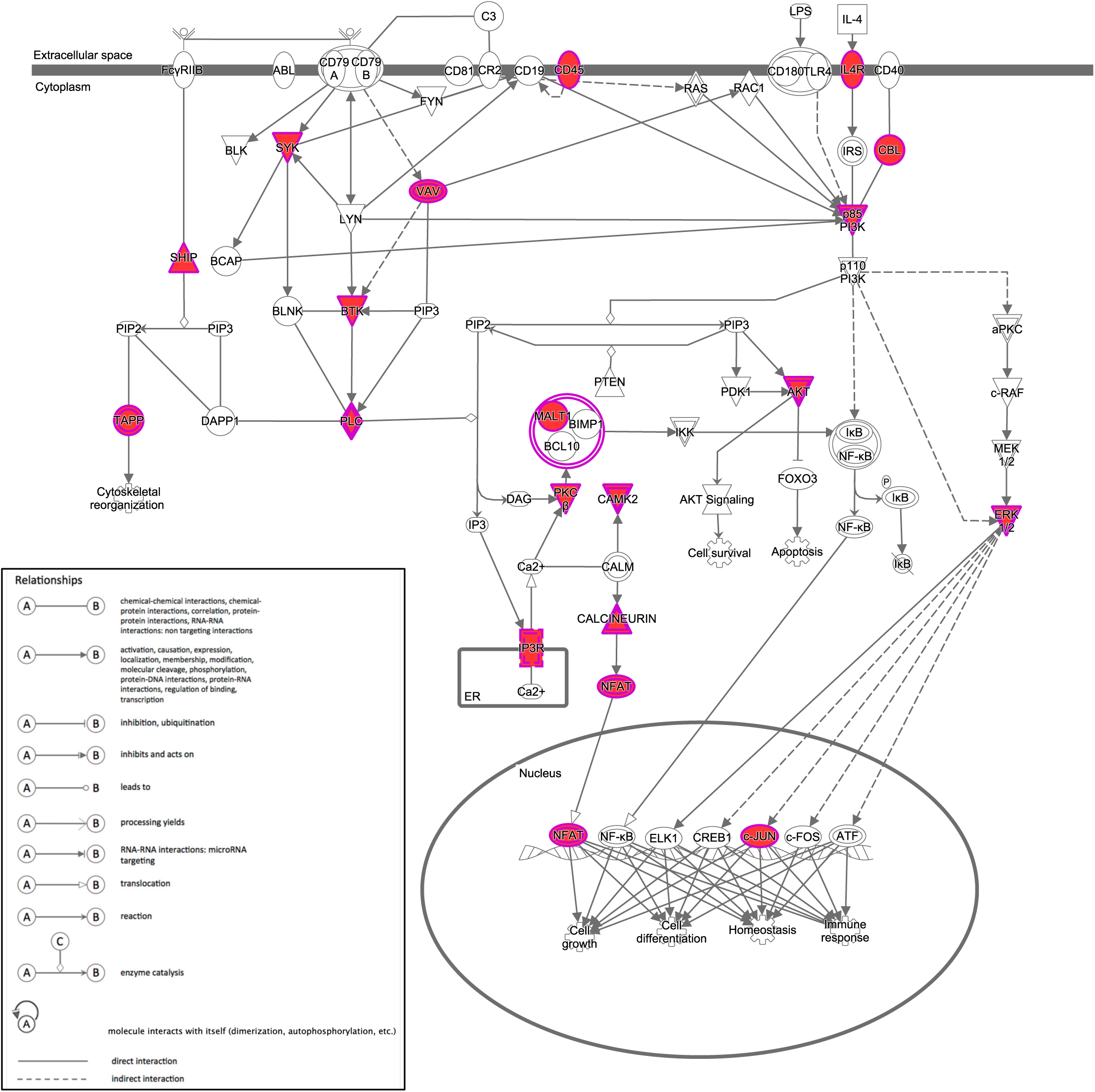
PI3K signaling in B cells is enriched in the recovering group. IPA canonical pathway analysis of genes significantly upregulated in B cells from recovering vs mild and severe COVID-19 patients. Analysis of the 539 DEGs upregulated (red) only in the recovering group shows increased activation of CD79 signaling compared to mild and severe groups.

**Supplemental Figure 9.**
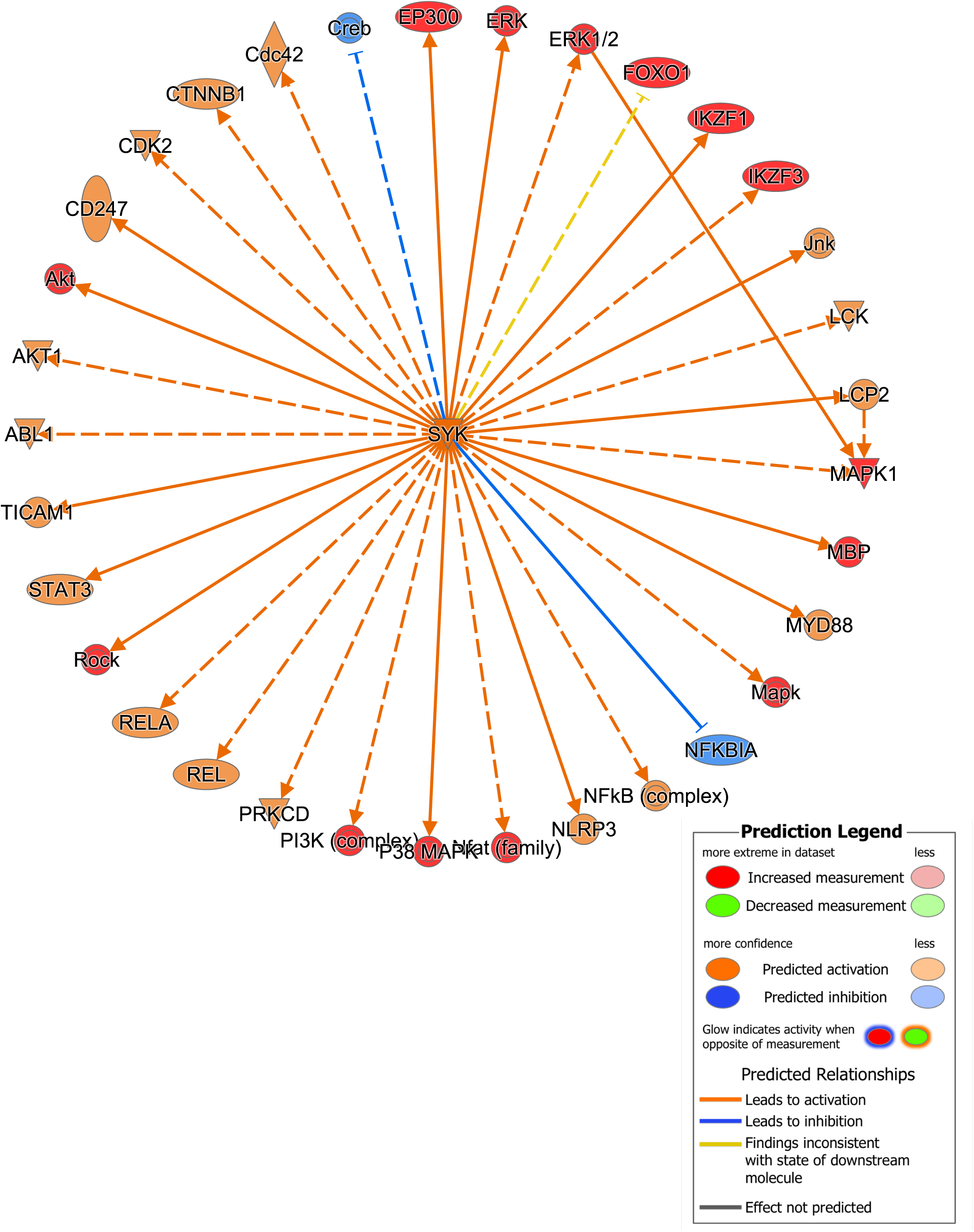
Increased B cell receptor signaling is the recovery group is regulated by SYK. IPA causal pathway analysis demonstrates that SYK is the primary upstream mediator of the upregulated pathways in B cells from recovering vs mild and severe groups.

**Supplemental Figure 10.**
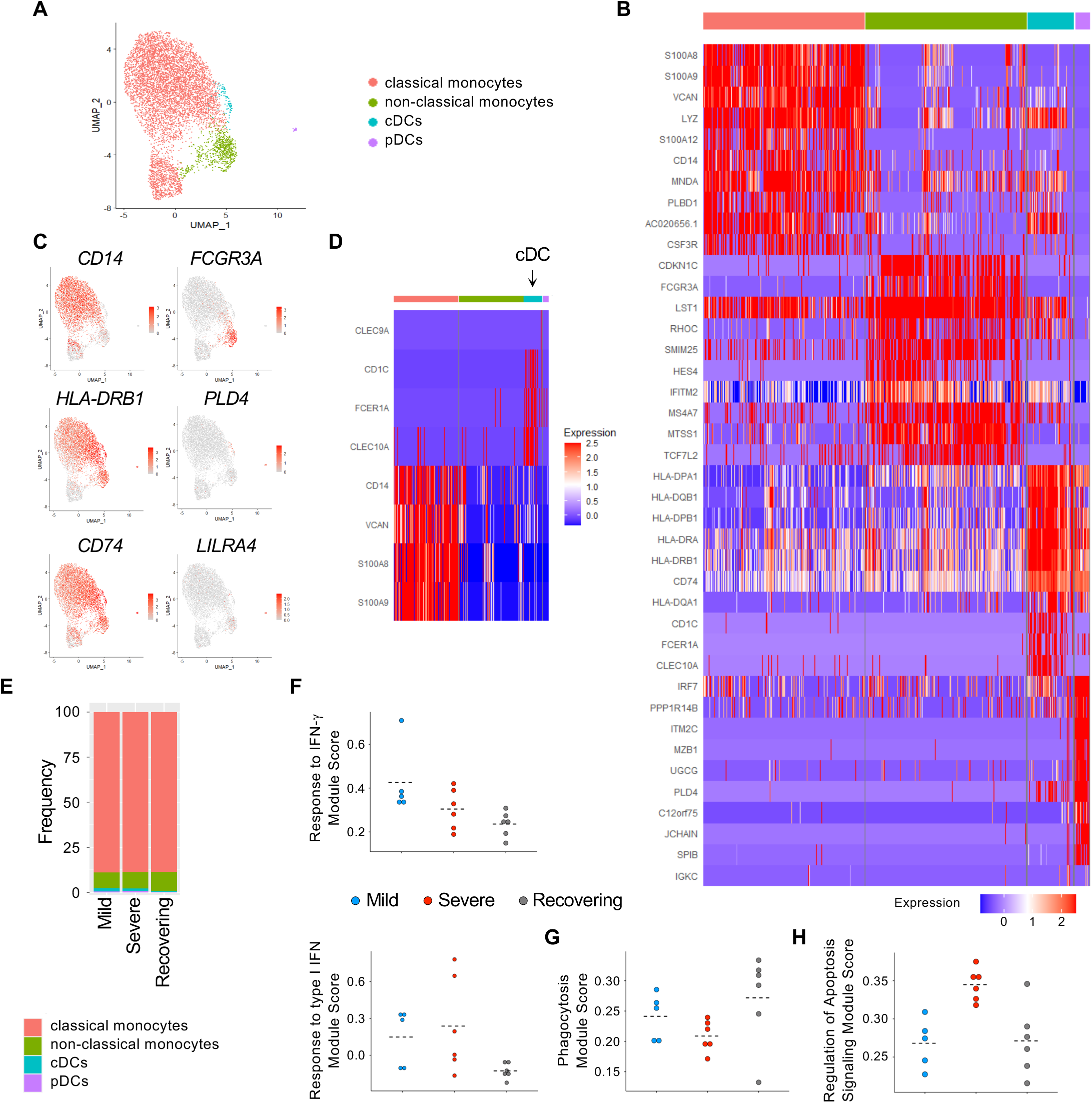
Identification of monocyte and DC subsets. A) UMAP of monocytes and DCs from all patients. B) Heatmap of genes differentially expressed by monocytes and DC subsets (downsampled). C) Monocyte and DC clusters were identified as classical monocytes (CD14), non-classical monocytes (FCGR3A), cDCs (HLA-DRB1, CD74) and pDCs (PLD4, LILRA4) cells. D) cDCs are predominantly DC2 cells (CD1C, FCER1A, CLEC10 with little/no monocyte gene expression). E) Proportions of monocyte and DC subsets in mild, severe and recovering patients. F-H) Mean module scores for individual patients.

**Supplemental Figure 11.**
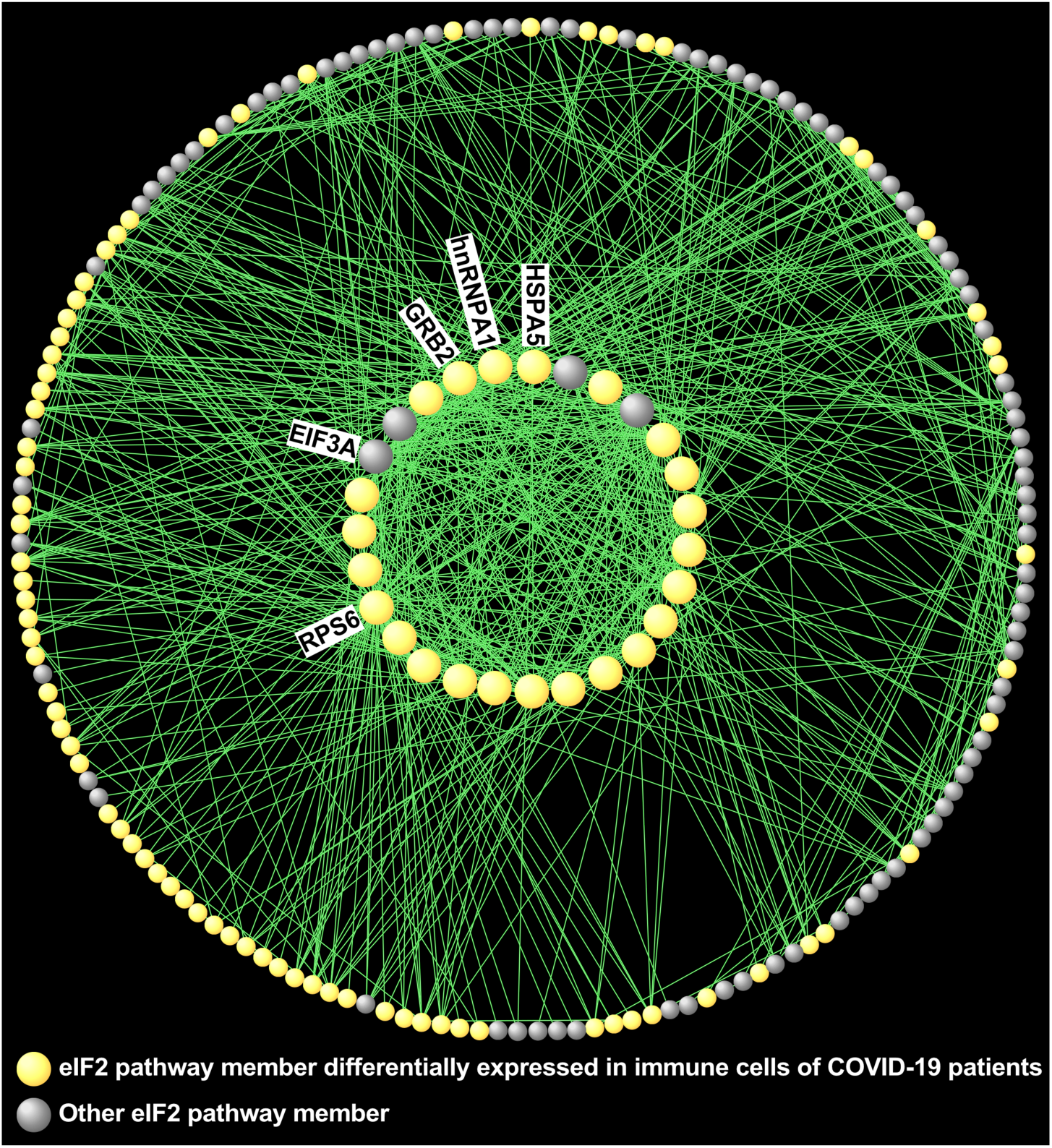
The eIF2 signaling pathway is differentially expressed in immune cells from COVID-19 patients. Gene product interaction network analysis of the eIF2 pathway, which is down-regulated in lymphocytes, but not not myeloid cells, in the severe vs mild group.

